# Right atrial dilatation associates with conduction velocity, incidence of arrhythmias and clinical outcomes in pulmonary hypertension

**DOI:** 10.1101/2025.02.13.25321996

**Authors:** S Ashwin Reddy, Jennifer T Middleton, Eckart MDD De Bie, Sarah L Nethercott, Gary J Polwarth, Katherine Bullock, Kiran Hk Patel, Nikesh Bajaj, Ben K Statton, Syeda Anum Zahra, Mahmoud Ehnesh, Caroline H Roney, Fu Siong Ng, David G Kiely, Andrew J Swift, Colin Church, Gerry Coghlan, Stefan Gräf, Allan Lawrie, James Lordan, Robert V Mackenzie Ross, Martin R Wilkins, S John Wort, Dolores Taboada, Katherine Bunclark, John E Cannon, Karen S Sheares, Joanna Pepke-Zaba, National Cohort Study of Idiopathic and Heritable PAH Collaboration, UniPHy Clinical Trials Network, Claire A Martin, Alex MK Rothman, Mark R Toshner

**Author notes:** **Corresponding author** Name: Mark Toshner, Postal Address: VPD Heart & Lung Research Institute, University of Cambridge. Joint senior author (Claire A Martin, Alex MK Rothman, Mark R Toshner).

## Abstract

**Introduction:** The mechanism, frequency and clinical impact of arrhythmias in pulmonary hypertension (PH) are unclear. We sought to clarify the electrophysiological mechanism in PH and correlate with arrhythmia incidence and clinical outcomes.

**Methods:** Invasive (n=10) and non-invasive (n=30) electrophysiological mapping techniques determined myocardial conduction velocity, scar burden and atrioventricular (AV) nodal refractoriness, which were then related to cardiac structure and function.

Implantable cardiac monitors (ICMs) were implanted into 80 patients with pulmonary arterial hypertension (PAH). Arrhythmia and clinical worsening episodes were prospectively assessed over 186 patient-years follow-up, and features associated with arrhythmia incidence were identified. Two cohorts encompassing all causes of PH (n=564 and n=3348) were interrogated to assess the relationship between defined arrhythmia risk variables and outcomes.

**Results:** Right atrial (RA) dilatation was associated with slower conduction velocity (R=0.52, p=0.01) and more prolonged AV nodal refractoriness (R=0.82, p=0.03). In the ICM study, 79 arrhythmia events were noted in 40% of participants. Only RA size was independently associated with significant arrhythmia (HR 1.03, 95% CI 1.01-1.06, p=0.01). Arrhythmia incidence associated with clinical worsening, and 20% of patients received ICM-uncovered targeted arrhythmia treatment. 69% of patients with treated arrhythmia were asymptomatic (median time-to-arrhythmia-detection 7 months). In both longitudinal PH cohorts, right atrial (RA) size was associated with worse survival independent of RA pressure and pulmonary vascular resistance.

**Interpretation:** The dominant mechanism associated with disordered cardiac electrophysiology in pulmonary hypertension patients is RA enlargement. RA size was the only significant variable associated with longitudinal arrhythmias in PAH and in two large prospective cohorts with representative populations of all causes of pulmonary hypertension, RA size is predictive of outcomes independent of pulmonary vascular resistance. Randomised trials are needed to clarify if screening and treating arrhythmias in pulmonary hypertension improves outcomes.

## Introduction

Adverse remodelling of the right heart in response to raised afterload, characteristic of pulmonary hypertension (PH), runs a pathological spectrum. The majority of work in this area has focussed on group 1 pulmonary arterial hypertension (PAH) where myocardial hypertrophy progresses to dilatation and subsequently patchy fibrosis of the right ventricle (RV) and further upstream, the right atrium (RA). This, allied to the high adrenergic tone and altered ion channel expression commonly associated with PAH, renders patients vulnerable to arrhythmias. In addition, disruption of atrioventricular and interventricular conduction is seen to occur at a higher frequency in idiopathic PAH (IPAH) patients compared to the general population^1^. More detailed characterisation of the electrophysiological substrate in pulmonary hypertension may be useful to identify pathophysiological features common to arrhythmic patients and relevant to all causes of pulmonary hypertension, which imprecise estimates currently suggest affects 1% of the global population^2^. The wider and more common global causes of pulmonary hypertension are a significant hidden public health unmet need.

Previous observational studies have sought to describe the incidence of arrhythmia in patients with PAH. The earliest of these were retrospective, single-centre analyses with heterogenous groups of PH patients (including those with PH secondary to left heart disease, congenital heart disease and CTEPH) ^3,4^, though subsequent prospective studies have corroborated the high rates of atrial arrhythmia reported in these early studies. Olsson et al. found a cumulative incidence of atrial flutter or fibrillation of 25.1% over 5 years in a population of 157 IPAH and 82 CTEPH patients^5^, whilst Wen et al. described a cumulative arrhythmia incidence of 15.8% over 6 years in 280 IPAH patients^6^. This contrasts with an incidence of 0.1% per year in the general population aged 55-60 years of age and a prevalence of 0.23%^7^. No study as yet has described the incidence of sustained ventricular arrhythmias or bradyarrhythmia in PAH.

The presence of either arrhythmias or conduction disease is associated with adverse outcomes in IPAH^1,3,5,6,8^, yet despite the clinical importance of prompt arrhythmia detection there is a paucity of studies investigating the arrhythmia incidence in any PH patients using long-term rhythm surveillance strategies^9^. Much of our current understanding of arrhythmia occurrence in this population is derived from observational studies using short-duration electrocardiographic monitoring detailed above ^3,5,6,10,11^, which means the true incidence is likely underestimated. Better understanding of arrhythmia burden and its clinical impact will allow identification of modifiable phenotypic risk-factors, aid risk-stratification, and inform patient selection for longer-term monitoring.

We sought to describe the electrophysiological and endotypic characteristics of the right heart and describe their relationship with respect to anatomical and structural parameters in patients with pulmonary hypertension using both invasive and non-invasive electroanatomical mapping methods, We then used minimally-invasive long-term arrhythmia monitoring in a prospective cohort of IPAH patients using implantable cardiac monitors (ICMs) to identify structural, functional and haemodynamic factors that associate with arrhythmia occurrence and clinical outcomes. Finally we validate our findings in two large prospective cohort studies of IPAH and all cause PH.

## Methods

### Arrhythmia substrate mapping

Adult patients with an established diagnosis of PAH/CTEPH, according to international criteria^2^, undergoing ablation of cardiac arrhythmia between 6/2019 and 6/2023 were eligible for inclusion into the invasive mapping arm of the study using the Rhythmia HDx™ mapping system (Boston Scientific, USA). Activation and bipolar voltage maps were collected simultaneously. Scar was defined as a local bipolar voltage of <0.5mV^12^. Following restoration of sinus rhythm, atrioventricular nodal effective refractory period (AVNERP) was measured with an 8-beat pacing drive train from the CS at a CL of 600ms followed by an extra stimulus (S2) starting at a coupling interval of 400ms and decreasing in 10ms increments down to a lowest of 200ms. The ERP was defined as the longest S2 which failed to propagate from atrium to ventricle. The RA electroanatomical maps were exported to Matlab for offline post-processing, involving calculation of RA volume, conduction velocity and scar burden-defined as percentage of scar relative to healthy myocardium. Conduction velocity was calculated using the cosine method assuming a planar wavefront propagation, as previously described^13^. Further details of the endocardial mapping procedure can be found in the supplementary appendix.

Thirty adult patients with an established diagnosis of IPAH who were scheduled to undergo cardiac magnetic resonance imaging (cMRI) on clinical grounds were invited to participate in the non-invasive mapping arm of the study using electrocardiographic imaging (ECGi), regardless of the prior history of cardiac arrhythmia. ECGi in conjunction with contemporaneously-acquired heart-torso geometry provides non-invasive electroanatomical assessment of the epicardium, and has been previously described^14^. Details of ECGi and cMRI protocols can be found in the supplementary appendix. The atrial and ventricular conduction characteristics of an age and sex-matched cohort of study patients were compared to a group of obese (n=16) and lean (n=16) patients from previously-published literature^15^.

The study was approved by the Cambridge East Research Ethics Committee (reference 20/EE/0177). All patients gave written informed consent before the study.

### Arrhythmia phenotyping and outcomes-National Cohort Study

80 patients were enrolled in the prospective study of arrhythmia phenotyping in PAH. This is a sub-study of the United Kingdom (UK) National Cohort Study of Idiopathic and Heritable Pulmonary Arterial Hypertension (ClinicalTrials.gov Identifier: NCT01907295), inclusion criteria for which are a current diagnosis of idiopathic, anorexigen-induced or heritable PAH. All patients enrolled in the National Cohort Study were eligible to participate and no additional arrhythmia-specific inclusion or exclusion criteria were applied for enrolment to the ICM sub-study. Patients were sequentially approached during clinical follow-up and enrolled into the study between October 2019 and September 2021 across two UK PH centres: Royal Papworth Hospital (RPH), Cambridge and Sheffield University Teaching Hospital, Sheffield. Demographic, arrhythmia symptom (defined as palpitations or syncope/presyncope), cardiac imaging, serological, pulmonary haemodynamic and functional data were collated for each patient as close to the implant as possible and at follow-up intervals as per local policy.

Each patient was implanted with a Reveal LINQ™ (Medtronic, Dublin, Ireland) subcutaneous implantable loop recorder (ILR) in the standard left parasternal position. Further detail regarding the device arrhythmia detection mechanisms can be found in the supplemental appendix. Participants were provided with a remote monitoring system (Carelink™). The electrograms of all arrhythmia events were manually reviewed by two cardiac electrophysiologists, and a diagnosis was only accepted when manual analysis confirmed the ILR classification.

All study patients were prospectively followed from implant to the census date (1^st^ March 2023), during which arrhythmia episodes, treatment interventions and clinical worsening events (defined as a composite of death, transplantation or unplanned hospitalisation due to PH deterioration) were logged.

Comparative analysis of baseline variables was undertaken to investigate features associated with arrhythmia occurrence. In patients who experienced significant arrhythmias over the study period multivariate analysis was carried out to discover independently associated clinical indicators. ‘Significant arrhythmias’ were defined as per established guidelines as any arrhythmia requiring pharmacological or invasive treatment, ventricular tachycardia (VT) or supraventricular tachycardia (SVT) lasting >30 seconds (‘sustained’ VT or SVT) ^16,17^ or asymptomatic pauses of over 3 seconds^18^.

A comparator group was created consisting of age and sex-matched patients without PAH who had ICMs implanted for established clinical indications^19^ over the same time frame. The event rate was compared between this and the PAH groups. The sub-study was approved by the UK National Research Ethics Service (13/EE/0203).

### Long-term outcomes-National Cohort study and ASPIRE registry

The association of independent arrhythmia risk factors, identified in the phenotyping arm of the study, with long-term mortality was estimated in a larger cohort of PAH patients through interrogation of the whole National Cohort Study of Idiopathic and Heritable Pulmonary Arterial Hypertension. Patients with at least one transthoracic echocardiogram (TTE) in the first 10 years post-diagnosis (n=564) enrolled in the National Cohort Study (NCT01907295). RAA was defined as ‘small’ if <22.8 cm^2^ and ‘large’ if >22.8cm^2^. Where multiple TTEs were recorded in the first 10 years post-diagnosis, median right atrial size was used. Findings in the National Cohort Study were then validated in the Assessing the Severity of Pulmonary Hypertension In a Pulmonary Hypertension REferral Centre (ASPIRE) cohort^20^. Ethical approval for ASPIRE was granted by the Institutional Review Board and approved by the National Research Ethics Service (16/YH/0352 subsequently 22/EE/0011). Patients diagnosed at >18 years with group 1 (n=1102), group 2 (n=717), group 3 (n=642), group 4 (n=788) or group 5 (n=99) PH were assessed for survival differences based on RAA at 10 years post diagnosis. The maximal RAA on a 4-chamber cardiac magnetic resonance imaging (cMRI) was used to define large (>22.8 cm2) or small (<22.8 cm2) RAAs. If multiple cMRIs were performed within 10 years of diagnosis, the median RAA was used.

### Statistical analysis

Continuous variables are presented as mean ± standard deviation or median [interquartile range], and categorical data as counts or percentages. Comparisons of parametric continuous data were performed using Student’s t-tests and analysis of variance (ANOVA), whilst categorical data were compared using the χ2 test. Comparisons of non-parametric data were performed using the Mann-Whitney-U test, or Kruskal-Wallis calculation for multiple-group testing. Significance was corrected for multiple-hypothesis testing using the false discovery rate (FDR) method (Benjamini-Hochberg adjustment). Survival was estimated using Kaplan-Meier analyses, with the log-rank test used to estimate significance. To determine independent risk factors for clinically significant arrhythmia occurrence, initial exploratory variables found to influence arrhythmia incidence with a P-value of <0.1 were used to generate a Cox proportional-hazards multivariate model. A two-tailed probability level of <0.05 was considered significant. Receiver operating characteristic (ROC) analysis was used to validate the predictive ability of significant variables, with the optimal cut-off point determined using the Youden index. The statistical strength and significance of correlation between structural and electrophysiological variables in the mapping arm of the study was determined by Spearman’s rank correlation coefficient. Statistical analysis was carried out with the R program (https://www.r-project.org).

Survival differences were assessed with a log-rank test for Kaplan-Meier curves with right censoring. Cox-proportional hazard models were generated, adjusting for age, sex, Pulmonary Vascular Resistance (PVR), mean Right Atrial Pressure (mRAP), and in the ASPIRE MR dataset additionally for Right Ventricular Ejection Fraction (RVEF) or Right Ventricular Systolic Volume Index (RVSV-I). Differences were assessed using a log-rank test. Previous studies demonstrate good correlation in atrial size estimates between echo and cMRI^21^.

## Results

### Arrhythmia substrate mapping

The baseline characteristics of both invasive and non-invasive cohorts are described in table S4, while table S5 describes the demographic and procedural details of patients undergoing invasive mapping. 30 PAH patients underwent non-invasive mapping with ECGi over the study period. Of these, ventricular mapping information was unusable for 10 patients and atrial information for 6 patients due to technical issues arising from data acquisition. MRI anatomical and strain data was performed for all patients. T1 mapping was unavailable for 9 patients as the necessary sequences were not performed (figure S1).

### Invasive assessment of cardiac conduction

Ten patients with PAH or CTEPH underwent invasive mapping and ablation for cardiac arrhythmia, all of which were for typical or atypical atrial flutter. Of these, six had an underlying diagnosis of CTEPH, two idiopathic or hereditary PAH and two PAH secondary to congenital heart disease (one closed ventricular septal defect, one open atrial septal defect). Conduction velocity (figure 1) and scar burden data was calculated for all ten cases.

**Figure 1:**
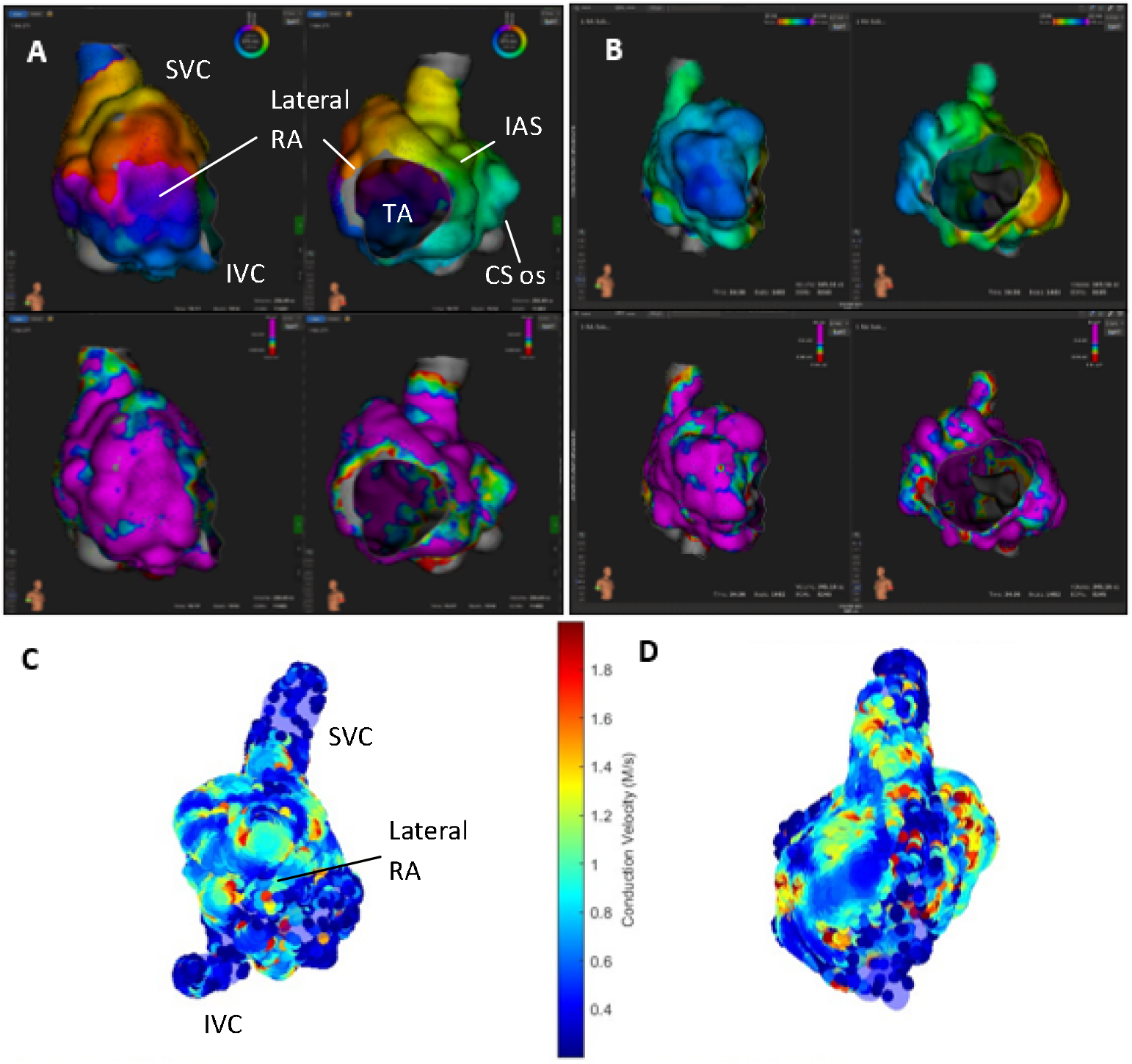
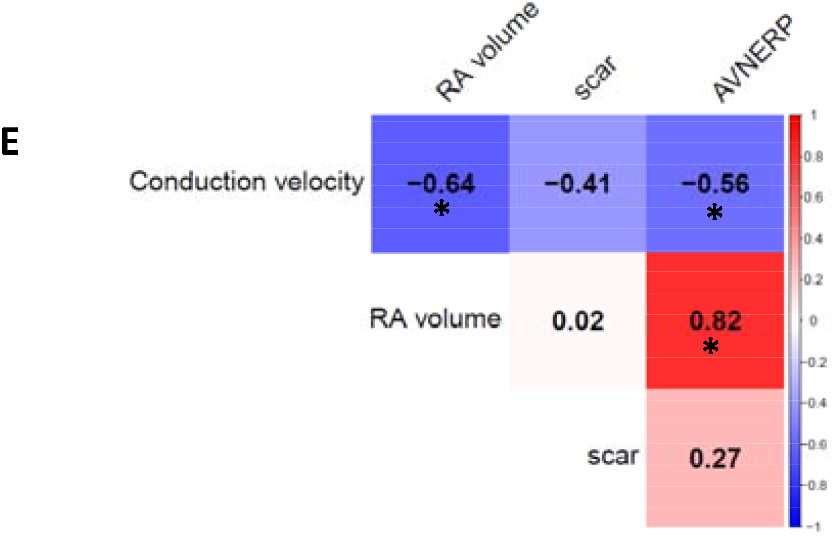
panels A and B-activation (top) and voltage (bottom) maps of the right atrium in right anterior oblique and left anterior oblique views in 2 patients undergoing invasive electrophysiology study and ablation. Panel A depicts a clockwise cavotricuspid isthmus-dependent flutter whilst panel B depicts a focal atrial tachycardia originating from the coronary sinus os. Panels C and D-atrial conduction velocity maps for the same patients. Panel E-heat map showing relationship between right atrial volume, atrioventricular nodal conduction, right atrial scar burden and right atrial conduction velocity on invasive catheter-based mapping. * denotes statistical significance (p <0.05). IVC= superior vena cava; IVC= inferior vena cava; TA= tricuspid annulus; RA= right atrium; CS= coronary sinus; AVNERP= atrioventricular nodal effective refractory period.

Nine of the patients underwent cavotricuspid isthmus (CTI) ablation for atrial flutter (including one for clockwise flutter), of whom two also had ablation for a second arrhythmia during the same procedure (one focal atrial tachycardia arising from the coronary sinus os and one pulmonary vein isolation for AF). The other patient underwent ablation of the lateral RA free wall for an atypical flutter (figure 1). Eight were in arrhythmia at the time of procedure. In one patient temporary pacing and overnight monitoring were required due to sluggish sinus node recovery post-ablation and arrhythmia termination; this patient was stable and discharged home the following day. There were no reported complications in the other cases. Atrial flutter did not recur in any patient by the time of study censor, though three experienced AF in early follow-up requiring re-initiation of antiarrhythmic medication. Clinical worsening events were noted in 3 ablation patients: two patients died, attributable to PH worsening, and one patient required PH-therapy uptitration due to clinical deterioration (table S5).

RA volume strongly and significantly positively correlates with AVNERP (R=0.82, p=0.03) and negatively with atrial conduction velocity (R=-0.64, p=0.05), but does not correlate with scar burden (R=0.02, p=0.95) (figure 1E).

Whilst atrial conduction velocity was negatively correlated with atrial scar burden this was not found to be statistically significant in this sample (R=-0.41, p=0.24) (figure 1E).

### Non-invasive assessment of cardiac conduction

Mean atrial activation time (AAT) was 75±14ms in the study group (n=23), ventricular activation time (VAT) 45±20ms (n=20), ventricular repolarisation time (VRT) 125±41ms, and ventricular activation-recovery interval corrected for RR interval (ARIc) was 286±26ms.

When an age and sex-matched selection of study patients (n=16) were compared to a previously published cohort of 16 disease-control obese patients and 16 healthy controls, AAT was significantly longer in PAH patients (72±17ms) than lean (46±12ms, p=0.001) but not obese (62±15ms, p=0.16) comparators. VAT similarly was significantly longer in PAH (41±13ms) than lean (31±6ms, p=0.009) but not obese (34±5, p=0.09) participants, and VRT was 127±51ms in the study cohort compared to 105±16ms in the obese (p=0.19) and 87±25ms in the lean (p=0.007) cohorts. Ventricular ARIc however was significantly longer in the PAH group (292±30ms) than both obese (247±19ms, p=0.0001) and lean (246±25ms, p=0.001) participants (table S6).

RA dilatation was associated with slower atrial activation (thus slower conduction) (R=0.51, p=0.01), and more prolonged repolarisation (R=0.40, p=0.02). There was no significant observed association between RV indexed volume and ventricular activation time (R=0.26, p=0.27) or activation recovery interval (R=0.07, p=0.78), but ventricular repolarisation significantly moderately inversely correlated with RV size (R=-0.50, p=0.02) (figure 2A).

**Figure 2:**
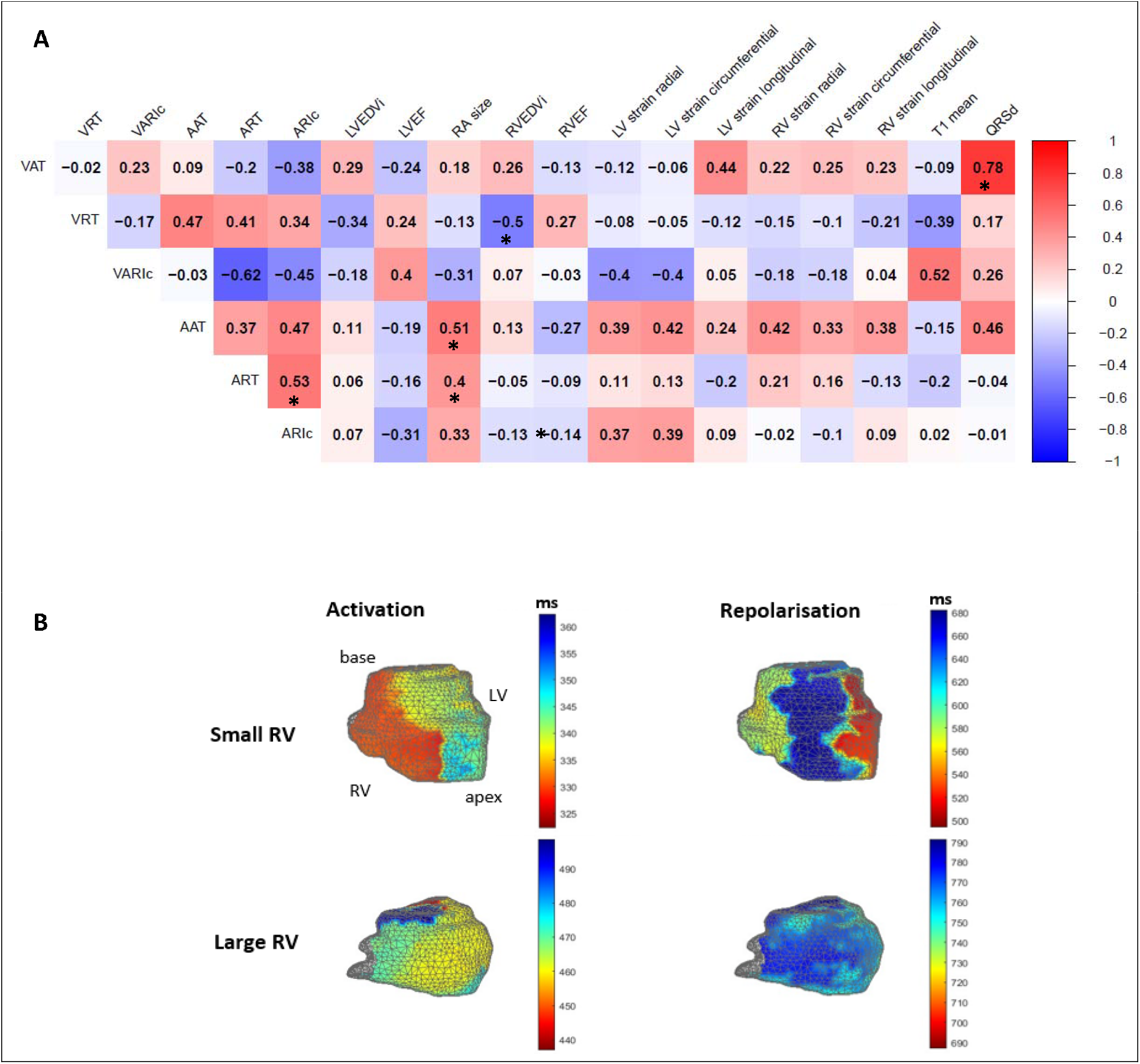
A-heat map showing correlation coefficients between structural, functional and electroanatomical variables derived from non-invasive mapping with ECGi and cardiac MRI. * denotes statistical significance (p <0.05). B-ventricular activation and repolarisation maps showing difference in gradients between larger and smaller ventricles. AAT= atrial activation time; ART= atrial repolarisation time; VAT= ventricular activation time; VRT= ventricular repolarisation time; ARIc= corrected activation-recovery interval; RA=right atrium; RV= right ventricle; LV= left ventricle; EF= ejection fraction; EDVi= indexed end diastolic volume

Ventricular activation time did not correlate with RV contractile function, whether assessed by RV ejection fraction (R=0.18, p=0.45) or strain (figure 2A).

RV scar burden, as defined by T1 mapping, was not found to correlate either with RV indexed volume (R=0.29, p=0.20) or ventricular activation time (R=0.005, p=0.99) (figure S4).

Scar burden moderately inversely correlates with RV ejection fraction (R= -0.58, p=0.006) and longitudinal strain (R=0.51, p=0.04) but is not significantly related to radial or circumferential strain (figure S5).

### Longitudinal ICM arrhythmia monitoring

Over the study period ICMs were implanted in 80 patients. The average age of the cohort was 53 ± 15 years and 23.8% were male. 38.8% of the study cohort had reported arrhythmia symptoms before, and a confirmed diagnosis of paroxysmal or permanent arrhythmia had previously been made in 15%. 12 patients additionally had previously undergone Holter monitoring, none of which had revealed acute arrhythmia requiring alteration of treatment. 19 (23.8%) patients were on antiarrhythmic therapy at time of ICM implant, which includes six out of 10 patients on non-dihydropyridine calcium channel blockers for treatment of vasoreactive PAH (all six on diltiazem, median daily dose 600mg). Baseline demographic and clinical features of the 80 study patients at the time of ICM implant are shown in table S1 of the supplementary appendix. The comparator group comprised of 71 patients matched for age (53 ± 17 vs 53 ± 15 years, corr. p=0.98) and sex (23.9 vs 23.8% male, corr. p=0.98) in whom ICMs were implanted for clinical indications (palpitations, n=22; syncope n=46; cryptogenic stroke n=2; and ventricular arrhythmia, n=1) over the same time-period. The mean monitoring period was 848 ± 236 days in the study group and 850 ± 253 days in the comparator group (corr. p=0.98). There were no significant differences in rate of major co-morbidity between the IPAH and comparator groups (table S2).

79 arrhythmia events were noted in 32 (40.0%) study patients over the monitoring period vs 78 arrhythmia events in 29 (40.8%) patients in the comparator group. In 16 study patients these were clinically relevant, resulting in debilitating symptoms and/or changes in management. Tachycardia episodes were seen in 23 (28.8%) patients and bradycardic episodes in 11 (13.8%); 2 patients experienced both tachy- and bradycardic events. There was no significant difference between the proportion of patients who experienced any arrhythmia (corr. p=0.98), or specifically tachy-(28.8 vs 31.0%, corr. p=0.98) or bradyarrhythmia (13.8 vs 15.5%, corr. p=0.98) in the IPAH or comparator groups. There were, furthermore, no significant differences noted in the subtype of arrhythmia detected (table S2, figure 3).

**Figure 3:**
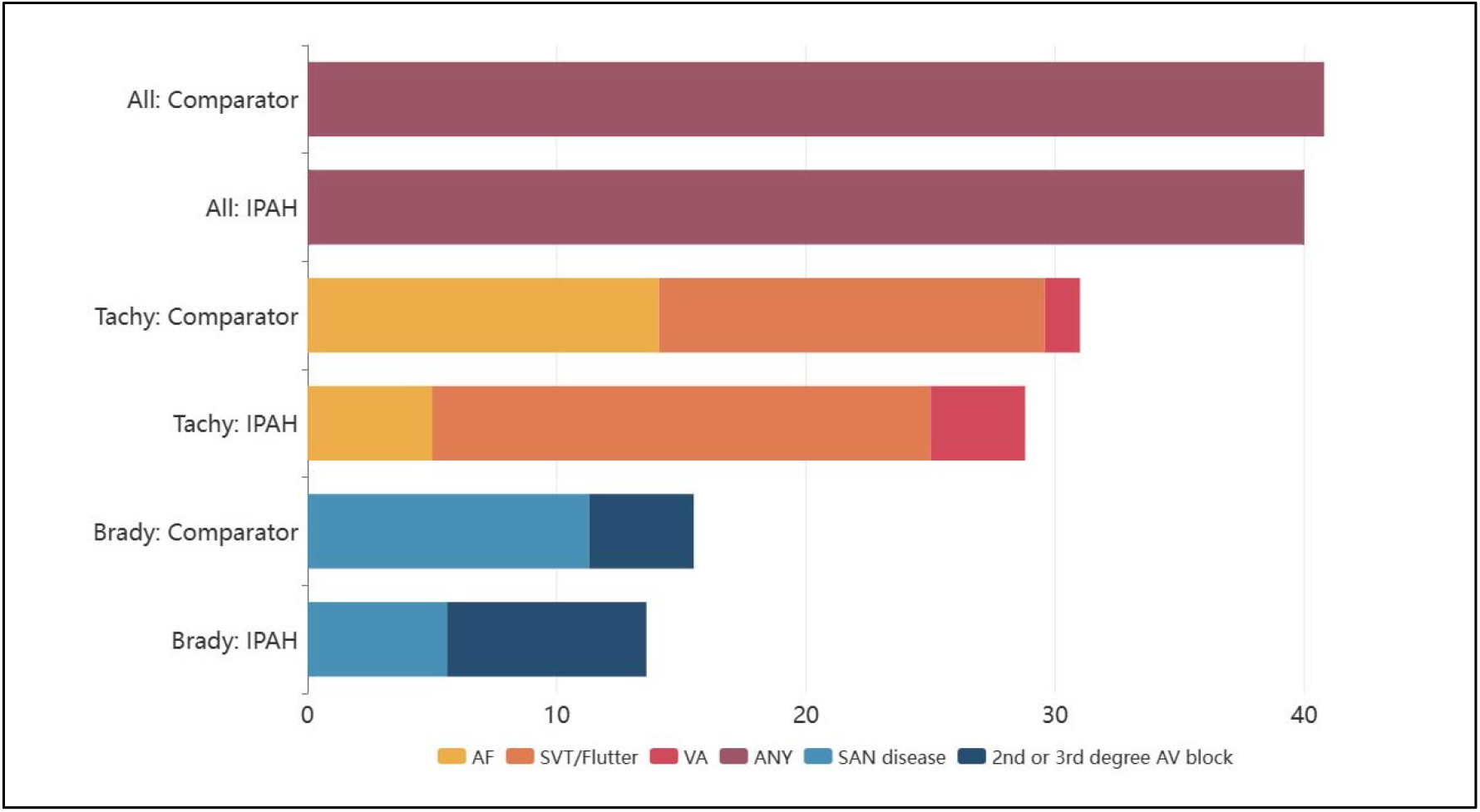
Percentage of IPAH and comparator patients with loop recorder-detected arrhythmia (left) and divided according to arrhythmia subtype (right). AF= atrial fibrillation; AFl= atrial flutter; AT= atrial tachycardia; SVT= supraventricular tachycardia; VA= ventricular arrhythmia; SAN= sinoatrial node; AV= atrioventricular.

### Factors associated with arrhythmia in PAH

Baseline characteristics of the study group, divided according to the presence or absence of arrhythmia, are shown in table 1. There was no significant difference in age, sex or BMI between arrhythmic and non-arrhythmic patients at time of ICM implant. PVR was higher in the tachyarrhythmic group compared to the bradyarrhythmic group, but not compared to the non-arrhythmic group; pulmonary haemodynamics were otherwise no different between arrhythmic and non-arrhythmic patients. Median time from RHC to ICM implant was 17 months. In unadjusted analyses, patients experiencing episodes of tachyarrhythmia had significantly higher NTproBNP levels at the time of ICM implant compared to both brady- and non-arrhythmic patients (491 [125-2019] vs 144 [63-786], p=0.04; vs 118 [76-352], between-group p= 0.02). A significantly greater proportion of patients with either brady-(36.4%) or tachyarrhythmia (34.8%) had 1st degree AV block or bundle branch block on their baseline ECG than those who remained arrhythmia-free (10.4%; between-group p=0.01). No differences remained significant after multiple test correction.

**Table 1:** Baseline demographic and clinical characteristics of the study cohort as a function of arrhythmia presence. Data presented as counts (%), mean ± standard deviation or median [interquartile range]. WHO= World Health Organisation; AV= atrioventricular; BBB= bundle branch block; RV= right ventricular; RA= right atrial; LV= left ventricular; LA= left atrial; EDV= end diastolic volume; ESV= end systolic volume; (i)= indexed to body surface area; PAP= pulmonary artery pressure; PCWP pulmonary capillary wedge pressure=; PVR= pulmonary vascular resistance; 6MW= six-minute walk. **‡ and * = between-group p value <0.05**.

|  | No<br>arrhythmia<br>N=48 | Tachy<br>N=23 | Brady<br>N=11 | P val | Corr.<br>P val |
| --- | --- | --- | --- | --- | --- |
| **Age** | 52.4 ± 15.2 | 51.0 ± 14.4 | 59.5 ± 14.9 | 0.28 | 0.54 |
| **Male sex** | 11 (22.9) | 4 (17.4) | 5 (45.5) | 0.19 | 0.47 |
| **Body mass index/kg.m<sup>-2</sup>** | 30.0 ± 5.2 | 29.2 ± 6.8 | 29.9 ± 4.4 | 0.84 | 0.95 |
| **Time from diagnosis/years** | 9.0 ± 5.2 | 8.8 ± 4.6 | 9.3 ± 4.6 | 0.96 | 0.98 |
| **Arrhythmia symptoms** | 6 (12.5) *‡ | 10 (43.4) * | 5 (45.5) ‡ | 0.07 | 0.32 |
| **WHO Functional Class** |  |  |  | 0.93 | 0.98 |
| I-II | 22 (45.8) | 10 (43.5) | 4 (36.4) |  |  |
| III | 19 (39.6) | 10 (43.5) | 6 (54.5) |  |  |
| IV | 7 (14.6) | 3 (13.0) | 1 (9.0) |  |  |
| **Baseline ECG** |  |  |  |  |  |
| Atrial fibrillation/flutter | 1 (2.1) | 3 (13.0) | 1 (9.1) | 0.18 | 0.47 |
| 1<sup>st</sup> degree AV block or BBB | 5 (10.4) *‡ | 8 (34.8) * | 4 (36.4) ‡ | 0.02 | 0.16 |
| Frontal plane axis deviation | 16 (33.3) | 9 (39.1) | 5 (45.5) | 0.72 | 0.95 |
| **Cardiac structure: MRI** | N=40 | N=18 | N=11 |  |  |
| RV EDV(i)/ml.m<sup>-2</sup> | 100.9 ± 38.6 | 96.7 ± 37.9 | 93.9 ± 26.8 | 0.83 | 0.95 |
| RV ESV(i)/ml.m<sup>-2</sup> | 61.4 ± 34.9 | 57.6 ± 33.0 | 52.1 ± 19.7 | 0.71 | 0.95 |
| RV ejection fraction/% | 41.8 ± 14.7 | 43.6 ± 13.9 | 44.4 ± 10.1 | 0.81 | 0.95 |
| LV EDV(i)/ml.m<sup>-2</sup> | 70.6 ± 16.2 | 72.4 ± 15.9 | 73.6 ± 14.4 | 0.84 | 0.95 |
| LV ESV(i)/ml.m<sup>-2</sup> | 29.9 ± 9.0 | 33.5 ± 10.0 | 32.4 ± 6.7 | 0.34 | 0.61 |
| LV ejection fraction/% | 63.7 ± 12.1 | 64.3 ± 9.7 | 59.8 ± 14.5 | 0.59 | 0.94 |
| RA area/cm<sup>2</sup> | 21.2 ± 4.4 ‡* | 25.9 ± 9.8 ‡ | 24.6 ± 8.9* | 0.02 | 0.16 |
| RA volume (indexed)/ml.m<sup>-2</sup> | 35.9 ± 11.3 ‡ | 50.7 ± 24.9 ‡ | 40.9 ± 18.8 | 0.01 | 0.16 |
| LA area/cm<sup>2</sup> | 21.7 ± 5.1 | 20.1 ± 5.1 | 23.6 ± 6.9 | 0.11 | 0.37 |
| **Pulmonary haemodynamics** |  |  |  |  |  |
| Systolic PAP/mmHg | 80 ± 24 | 87 ± 26 | 73 ± 23 | 0.26 | 0.54 |
| Diastolic PAP/mmHg | 31 ± 11 | 35 ± 11 | 28 ± 9 | 0.25 | 0.54 |
| Mean PAP/mmHg | 48 ± 14 | 54 ± 17 | 43 ± 15 | 0.11 | 0.37 |
| PCWP/mmHg | 11 ± 4 | 10 ± 3 | 11 ± 2 | 0.66 | 0.95 |
| PVR (Thermodilution)/dynes | 848 ± 541 | 1119 ± 700 ‡ | 571 ± 268 ‡ | 0.03 | 0.16 |
| RA pressure/mmHg | 9 ± 6 | 9 ± 5 | 9 ± 3 | 0.98 | 0.98 |
| Cardiac index | 2.2 ± 0.9 | 2.0 ± 0.7 | 2.6 ± 0.6 | 0.18 | 0.47 |
| **NTproBNP/pgml<sup>-1</sup>** | 118 [76-352]<br>* | 491 [125-2019] *‡ | 144 [63-786]<br>‡ | 0.03 | 0.16 |
| **6MW distance/metres** | 357 ± 218 | 381 ± 190 | 459 ± 183 | 0.37 | 0.62 |

Eleven out of 80 patients did not undergo cardiac MRI: (6 patients declined, 5 had implanted devices or instruments precluding routine scanning). Among the 69 patients who did undergo cardiac MRI, we found that right atrial size was significantly larger in arrhythmic patients, particularly those experiencing tachyarrhythmia (respectively for no arrhythmia vs. tachyarrhythmia vs bradyarrhythmia: RA area 21.2 ± 4.4 vs 25.9 ± 9.8 vs 24.6 ± 8.9cm^2^, between group p<0.05 between non-arrhythmic and arrhythmic; RA indexed volume 35.9 ± 11.3 vs 50.7 ± 24.9 vs 40.9 ± 18.8 ml.m^-2^, between group p<0.05 between non-arrhythmic and tachyarrhythmic). Other measures of right and left heart structure and function were similar between groups.

### Arrhythmia-associated outcomes and associated features

The presence of arrhythmia during the study period was significantly associated with time to clinical worsening events (figure 4A). Clinical worsening occurred in 5 (10.4%) non-arrhythmic patients (2 PH-related hospitalisation, 2 deaths, 1 transplant) and 9 (28.1%) arrhythmic patients (4 PH-related hospitalisations, 4 deaths, 1 transplant with subsequent death).

**Figure 4:**
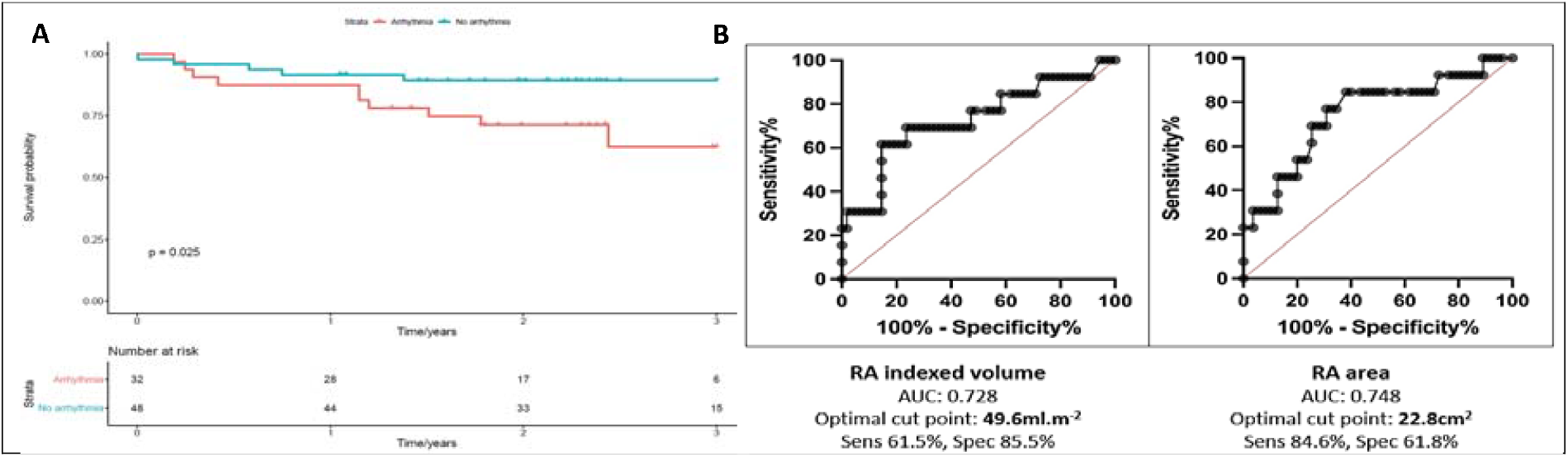
A-Survival to clinical worsening event (death, transplant, pulmonary hypertension-related hospitalisation) as a function of presence or absence of arrhythmia. B-Receiver-operator characteristic curves of indexed right atrial volume (left) and right atrial area (right) as predictor of significant arrhythmic events. RA= right atrial; AUC= area under curve.

Eight (16.7%) non-arrhythmic patients had improved their risk stratum by the end of the study, as defined by COMPERA 2.0 4-strata risk classification22 and seven (14.6%) ended up in higher risk groups or had died, whereas in patients experiencing arrhythmia four (12.5%) improved and six (18.8%) worsened (tachyarrhythmic patients: two (8.7%) improved and five (21.7%) worsened; bradyarrhythmic patients: three (27.3%) improved and one (9.1%) declined) (figure S2).

Sixteen (20.0%) patients had changes to treatment as a consequence of significant arrhythmia identified by ICM necessitating intervention, details of which can be found in the supplementary appendix. Eleven of these patients were asymptomatic with the detected arrhythmia. In total, 29 significant arrhythmic events occurred between one and 23 months of ICM implant. The time to first significant arrhythmia ranged from one to thirteen months (median= seven months).

Variables associated with arrhythmia with a significance value of ≤0.1 (symptoms, atrioventricular conduction disease, NTproBNP and right atrial size) were included in multivariate analysis. Right atrial size alone was independently associated with the occurrence of clinically significant arrhythmia (table S3). Separate analyses of the atrial area (related directly to volume) with symptoms, atrioventricular conduction disease and NTproBNP also demonstrated the same result (HR 1.03, 95% confidence interval 1.01-1.06, p=0.01). ROC curve analysis showed an area under the curve for indexed right atrial volume of 0.728 (p=0.01) and right atrial area of 0.748 (p=0.006) (figure 4B). The optimal cut point for indexed RA volume was 49.6ml/m^2^ (sensitivity 61.5%, specificity 85.5%), and for RA area was 22.8cm2 (sensitivity 84.6%, specificity 61.8%).

### Mortality associations in the National Cohort Study and ASPIRE registry

RA area was significantly associated with worse 10-year mortality. Using an RA area cut-off of 22.8cm2 to dichotomise patients, based on our findings described above, the mortality difference was highly statistically significant (log-rank p<0.0001) across two large cohorts of group 1 (figures 5A and S6) and all-cause PH (figure 5B). In the National Cohort study 5-year mortality was 74.9% and 82.5% for larger and smaller atrial area respectively.

**Figure 5:**
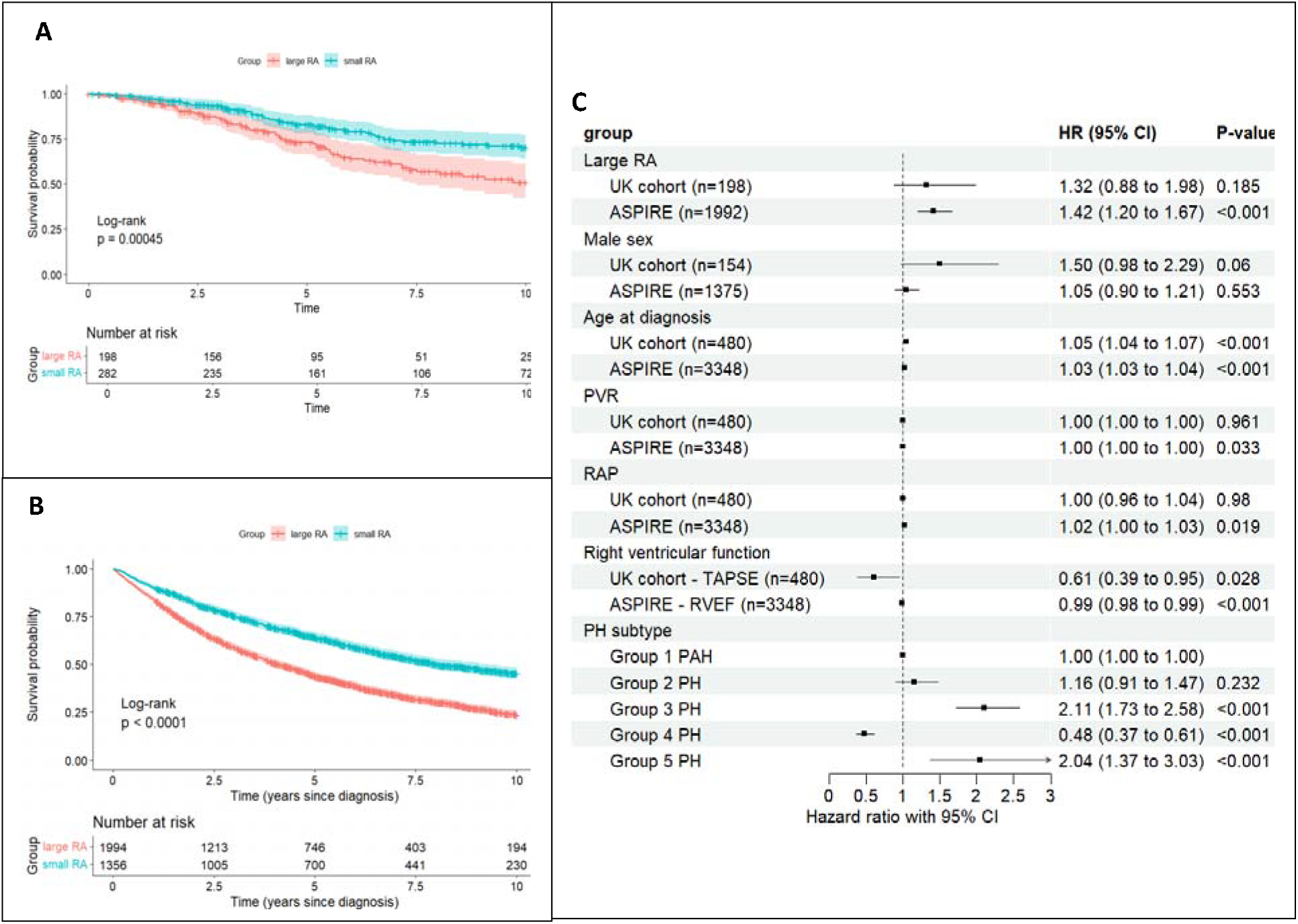
Kaplan-Meier analysis depicting 10-year survival of patients with (panel A) group 1 and (panel B) all-cause pulmonary hypertension as a function of right atrial size dichotomised according to area < or >22.8cm2. Panel C-Cox proportional hazard Forest plot depicting independence of association between indexed right atrial size, age, sex, pulmonary haemodynamics and right ventricular function to mortality in pulmonary arterial hypertension patients. RA= right atrial; BSA= body surface area; RAP= right atrial pressure; PAWP= pulmonary arterial wedge pressure; PAP= pulmonary artery pressure; APSE= annular plane systolic excursion

RA area of >22.8cm2 remained significantly associated with survival in both group 1 and all-cause PH in the ASPIRE registry even when age, sex, RV function and pulmonary haemodynamics were accounted for on multivariable analysis (all-cause PH-HR 1.42, 95% CI 1.20-1.67, p=<0.001) (figure 5C, table S8). The same relationship was observed for patients in the National Cohort Study but, with smaller patient numbers, this was not found to be statistically significant (figure 6C). In addition, RAA was a significant independent predictor of survival across all different individual PH groups (figure S6).

## Discussion

In prospective, multi-centre studies we demonstrate that patients with IPAH have a high incidence of arrhythmia episodes. Right atrial dimensions are associated with mechanistically convincing changes for arrhythmia-generative features and are associated with outcomes in all causes of PH independent of classical haemodynamics.

Using both non-invasive ECGi-based and invasive catheter-based electroanatomical mapping, we identify likely mechanisms whereby RA dilatation may mediate arrhythmogenesis: firstly by demonstrating slower RA conduction velocity with progressively dilating chamber size (thus predisposing to tachyarrhythmia), and secondly by determining that AV nodal refractoriness is strongly and significantly related to increasing RA chamber volume (thus predisposing to atrioventricular conduction disease). Ventricular conduction however does not appear to correlate with chamber size, function or scar burden. In addition, we show that RA and RV conduction is markedly slower in patients with PAH compared to healthy or obese patients without PAH.

The prognostic value of RA size is a well-described in PAH, and its importance for risk stratification is reflected by its inclusion in current international guidelines^2^. We validate this across all causes of PH and demonstrate that the association between RA size and mortality is independent of RAP, PVR and RVEF, strongly suggesting that the effect on mortality is not simply mediated by an association with increasing PVR or RV dysfunction. We also show that RA enlargement is independently associated with arrhythmias in this patient group and, furthermore, that PAH patients who experience arrhythmia have worse outcomes. We contend that a contribution of right atrial dilatation’s prognostic value is mediated through cardiac arrhythmogenicity. A major question now is whether reduction of RA size through aggressive management of preload or afterload may mitigate arrhythmia risk in PAH as it does mortality^23^ and whether aggressive anti-arrhythmic treatment will be of additional value.

Right atrial dilatation and maladaptive remodelling in PAH, resulting from upstream transmission of chronically elevated right ventricular afterload, leads to focal deposition of collagen types I and III between cardiomyocytes mediated largely by resident cardiac fibroblasts, by far the most abundant interstitial cells of the myocardium. This process contributes to electric uncoupling of the cardiomyocyte connections, leading to disrupted electric propagation^24^. Even in the absence of fibrotic change, mechanical stretch may alter the electrophysiological properties of the myocardium through alteration in myocardial cell-surface ion channel expression and intracellular calcium handling in response to distension^25^. Delayed conduction across the crista terminalis has been observed in patients with chronic right atrial stretch resulting from atrial septal defects^26^, and is only partially corrected on closure of the septal defect. In addition, acute atrial stretch results in slower atrial conduction velocity in human and mammal experimental models^27^ and is independent of load. The consequence of slowed conduction in dilated right atria is the potential for generation of re-entry circuits, as a slowly propagating conduction wavefront encounters adjacent healthy myocardium that is no longer refractory and excites it. This mechanism, first described by Mines in 1913, is a pre-requisite for many tachyarrhythmias and fibrillation. Atrial dilatation also strongly and significantly correlated with longer AV nodal refractoriness on invasive electrophysiology studies. This has been observed in clinical PAH cohorts and is associated with worse outcomes ^1,8,28^.

Previous studies have implicated atrial size, strain or load with arrhythmia occurrence ^5,6,29,30^. We likewise noted higher NTproBNP and more pronounced right atrial enlargement at time of ICM implant in patients who developed tachyarrhythmia, thus corroborating these findings. However, in contrast to some prior studies, we did not find any significant difference in invasive haemodynamic parameters or demographic factors between arrhythmic and non-arrhythmic patients. Further to this, we note that clinically important arrhythmia in particular was independently associated with larger RA area and volume, but not other traditional risk factors. This agrees with data on hospitalisation outcomes in arrhythmia and therefore is relevant to clinical worsening^30^. We acknowledge the temporal dyssnchrony between RA measurements, which were averaged over follow-up, and haemodynamic measures, which were taken at baseline, though we feel our conclusions are strengthened by other contemporaneous data corroborating these findings. The comparable arrhythmia rates to established indications for ICM implantation might controversially suggest that all IPAH patients should be considered for ICM screening, however a better targeted approach might take into account (or stratify for) right atrial structure/size. Our ROC analyses suggest that RA area >23cm^2^ or indexed volume >50ml.m^2^ serve as useful starting thresholds for validation. Our work strongly suggests that a more rigorous approach to ICM-based assessment should involve consideration of RA structure and not be restricted to symptomatology only.

The increased sensitivity of ICMs in detecting intermittent paroxysmal cardiac arrhythmias is now well-established. Both the STROKE-AF and CRYSTAL-AF trials reported AF detection rates of 12% using ICMs vs 2% with either 12-lead ECG, Holter monitoring or event recording after 12 months in patients with cryptogenic stroke who had no prior documented evidence of AF. In the RUP study of 50 subjects with unexplained palpitations randomly assigned to ICM or conventional monitoring, a diagnosis was obtained in 21% of the conventional strategy group and 73% of the ICM group (p<0.001). In a recent prospective study of 34 patients (24 PAH, 10 CTEPH) without prior arrhythmia diagnosis, Andersen et al. reported that 70 arrhythmia episodes were recorded in 38% of patients over 46 patient-years follow-up. The majority were short-lived and did not require further intervention^9^. It is noteworthy that in our study all significant arrhythmia events occurred at least one month after ICM implantation, thus none would have been detected using standard-of-care ambulatory monitoring. Six of the patients with significant arrhythmia had previously undergone 24-hour Holter monitoring without any acute dysrhythmia being detected.

## Limitations

The presence of arrhythmia in our studies correlated with both mortality and deterioration requiring hospitalisation. In the only other studies comparable to our work patients had more advanced disease; 79% of Olsson’s cohort and 65% of Wen’s were in WHO functional class III or IV (vs 56% of ours), whilst the average NTproBNP in Mercurio’s study was 2071 and 6-minute walk distance 327.6m (vs 164 and 370m respectively). ^7–9,45,4646^

The comparator group in the arrhythmia phenotyping study comprised of patients who had clinical indications for ICM implantation, and therefore arrhythmia episodes were more likely to be seen than would be expected in a group of unselected age- and sex-matched individuals. As there was no direct control group in our study, or any prior study using ICMs to our knowledge, we cannot determine the relative frequency of arrythmia in IPAH patients compared to healthy subjects. The mean follow-up duration of 2.3 years from time of implant may be too short to determine the full effects of short-lived subclinical arrhythmia on longer term outcomes.

## Conclusions

We have demonstrated that in PAH, RA dilatation was associated with slower conduction velocity, longer AV nodal refractoriness, and that RA dilatation is associated with increased frequency of treatable arrhythmia and clinical outcomes. We have validated in large prospective cohorts of all-cause PH that RA size is associated with worse mortality, independently of PVR and right ventricular function. We postulate that this may be at least partly mediated by increased arrhythmia incidence. Prospective studies are now required to clarify if more aggressive identification and treatment of arrhythmia and atrial dilatation across pulmonary hypertension classifications groups will improve outcomes.

## Data Availability

All data produced in the present study are available upon reasonable request to the authors

## Acknowledgements and Conflicts of Interest

We wish to acknowledge Boston Scientific and Medtronic as funding sources for this study.

Alex Rothman would like to acknowledge research funding from the Wellcome Trust (206632/Z/17/Z).

Fu Siong Ng would like to acknowledge research funding from the British Heart Foundation (RG/F/22/110078 and RE/24/130023).

Rob McKenzie-Ross, Robin Condliffe, David Kiely: honoraria from Janssen.

Claire Martin: honoraria and consulting fees from Medtronic, Boston Scientific, Biosense Webster. The ASPIRE registry is supported by the National Institute for Health and Care Research (NIHR) Sheffield Biomedical Research Centre (NIHR203321). The views expressed are those of the author(s) and not necessarily those of the NIHR or the Department of Health and Social Care.

## Collaborators

**National Cohort Study of Idiopathic and Heritable PAH Collaboration:**

**Freeman Hospital, Newcastle**

James Lordan, Alan Greenhalgh

**Golden Jubilee National Hospital, Glasgow**

Martin Johnson, Colin Church, Val Irvine

**Hammersmith Hospital, London**

Martin Wilkins, Luke Howard

**Royal Brompton Hospital, London**

John Wort

**Royal Free Hospital, London**

Gerry Coghlan, Tani Ngcozana

**Royal Hallamshire Hospital, Sheffield**

Allan Lawrie, David Kiely, Chloe Roddis, Stefan Roman

**Royal Papworth Hospital, Cambridge**

John Cannon, Karen Sheares, Dolores Taboada, Katherine Bunclark, Joseph Newman, Jean Gosbee, Emily Knightbridge, Karen Brookes

**Royal United Hospital, Bath**

Robert Mackenzie-Ross, Katie White, Zandile Maseko, Jay Suntharalingam

**University of Cambridge**

Nicholas Morrell, Stefan Graf, Carmen Treacy

**ASPIRE Consortium**

David G Kiely, Lisa Watson, Iain Armstrong, Catherine Billings, Athanasios Charalampopoulos, Robin Condliffe, Charlie Elliot, Abdul Hameed, Neil Hamilton, Judith Hurdman, Allan Lawrie, Robert Lewis, Smitha Rajaram, Alex Rothman, Andy Swift, Steven Wood, Roger Thompson, Jim Wild

