## Supplementary appendix for "Right atrial dilatation associates with conduction velocity, incidence of arrhythmias and clinical outcomes in pulmonary hypertension"

Supplementary material

[Figure](#_Toc170730435) S1	5

[Table](#_Toc170730436) S1	6

[Table](#_Toc170730435) S2	8

[Figure](#_Toc170730436) S2	9

[Figure](#_Toc170730435) S3	10

[Table](#_Toc170730436) S4	10

[Table](#_Toc170730435) S5	12

[Figure](#_Toc170730436) S4	14

Figure S5	14

[Table](#_Toc170730436) S6	14

[Table](#_Toc170730435) S7	15

[Figure](#_Toc170730436) S6	16

### **Methods**

#### **ILR arrhythmia detection**

The arrhythmia detection algorithm in the Reveal LINQ™ ILR continuously assesses R-R interval regularity. Sufficient R-R variability over a 2-minute period is automatically recognised by the device as an arrhythmia. Specificity is improved through more recent development of improved P-wave sensing, which allows better discrimination between atrial tachycardia/fibrillation (AT/AF) and sinus arrhythmia. Bradycardia and pause detection algorithms are based on the duration of the sensed R-R interval. Patients can, additionally, make manual recordings when experiencing arrhythmic symptoms. The device memory can store up to 27 min of ECG recordings from automatically detected arrhythmias and up to 30 min of ECG recordings from patient-activated episodes. Date, time and duration of episodes are recorded. When storage capacity is met, older electrograms are overwritten with newer ones.

The ILR automatically transmits the last 10 seconds of the 2-minute ECG segment for the longest episode of the day to the remote Carelink monitor via Bluetooth every day. In addition, patients were asked to perform regular manual transmissions of the full information on all episodes stored in memory. This ensured they were recorded before being overwritten.

#### **Invasive endocardial mapping**

##### **Ablation procedure**

Patients undergoing electrophysiology (EP) study +/- ablation for diagnosis and treatment of atrioventricular (AV) node-dependent arrhythmias discontinued antiarrhythmic drugs (AADs) 5 days prior to the procedure. For typical or atypical flutter ablation AADs were continued until the day of ablation as per local protocol.

Procedures were carried out in a fasted state under sedation or general anaesthesia administered by a cardiothoracic anaesthetist. Vascular access was obtained via the femoral vein under ultrasonographic guidance, and a steerable decapolar catheter was placed in the coronary sinus (CS) and quadripolar catheters in the His position and right ventricular (RV) apex as appropriate. High-density maps of right atrial flutters were created with the Rhythmia HDx™ mapping system using the 64-electrode steerable Intellamap Orion™ basket catheter (Boston Scientific, USA), whilst ablation was carried out using an Intellanav™ open-irrigated catheter with the aid of a steerable sheath.

#### **Non-invasive epicardial mapping**

##### **Electrocardiographic imaging (ECGi) protocol**

Flexible strips containing 256 electrodes (BioSemi, Amsterdam, Netherlands) were attached to the front and rear torso of each patient and 5 minutes of surface electrocardiographic recording obtained at a sampling frequency of 2048 Hz at rest in a semi-recumbent supine position. As the electrodes are not MRI-conditional these were replaced in identical positions by MRI-opaque location markers, following which the patient was carefully transferred to the MRI scanner to ensure the location markers were not translocated. A cardiac MRI (cMRI) was then performed to demonstrate the heart’s location within the thorax relative to the electrode markers (‘heart-torso geometry’).

Electrocardiographic and imaging data were exported for post-processing. Epicardial meshes and electrode locations on the torso were reconstructed using commercial software (Amira, ThermoFisher). Each electrode marker was manually labelled in sequence of the electrodes placed on the torso. For cardiac geometry, the atrial and ventricular perimeters were manually traced from a single static image at each slice location on each of the 94 coronal sections in the end-diastolic phase, with the valve planes delimiting the extent of each chamber.

Unipolar epicardial electrograms were reconstructed by solving the inverse problem of electrocardiography and were processed to compute electrical parameters of interest. Activation time (AT) was computed as the steepest negative time-derivative of voltage (-dV/dtmax) in the local QRS complex. Recovery time (RT) was computed as the time point of steepest positive time-derivative (dV/dtmax) during the T-wave ^19^. Activation-recovery intervals (ARI, a surrogate for local action potential duration) were computed as the difference between RT and AT.

##### **Cardiac MRI protocol**

Each patient underwent a cardiac MRI scan, immediately after replacing the electrodes strips with MRI-safe markers, in either a Siemens 1.5 or 3 Tesla scanner with gadolinium contrast enhancement. A respiratory-navigated Half-Fourier Acquisition Single-shot Turbo spin Echo (HASTE) sequence in the transverse plane anatomical sequence was used so the ECGi electrical and anatomical data could be merged.

Conventional cine imaging and, where clinically indicated, T1 mapping and advanced late gadolinium enhancement (LGE) imaging were conducted following the thorax scan and analysed offline. Cine imaging was used to measure volumetric and functional data. Volumes were acquired in end-expiration breath-hold. Contouring at end-systole and end-diastole was performed using cvi42 version 5.10.1 (Circle Cardiovascular Imaging Inc, Calgary, Alberta, Canada).

LGE images were acquired using a free-breathing 2D inversion-recovery sequence approximately 3-5 minutes after administration of a bolus injection of an intravenous gadolinium-based contrast agent (0.3 mmol/kg; Dotarem) given manually, immediately followed by a 15-20 ml saline flush. The inversion time (TI) was adjusted for optimal nulling of remote normal myocardium.

RV myocardial scar burden was performed using modified look-locker inversion recovery (MOLLI) T1 mapping (MyoMaps, Siemens Healthcare, Erlangen, Germany). T1-maps were acquired from vendor-provided product protocols. For the analysis of MOLLI T1 maps, the myocardium of the short axis slice acquired at baseline was contoured using dedicated software providing a single average myocardial T1 value per individual. T1 maps were described according to anatomical RV segments: anterior wall, posterior wall, free wall and septum.

Radial and longitudinal endocardial and wall strain were derived from long axis 2, 3 and 4-chamber views, and circumferential strain from the short axis view at the midpapillary level.

### **Results**

Flow diagram of patient cohorts (figure S1) and baseline demographic and clinical characteristics of the ILR substudy (table S1).

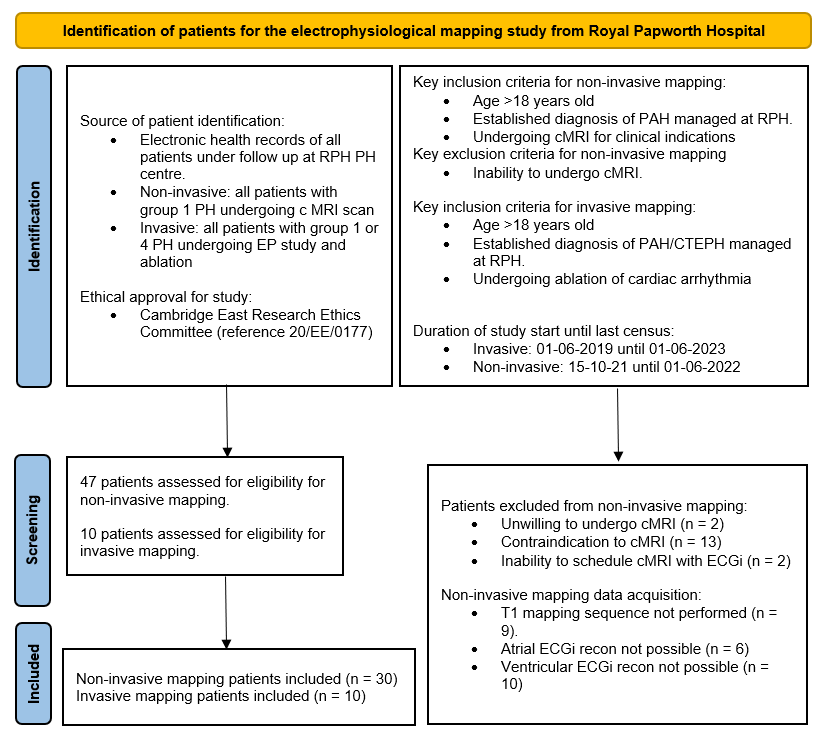

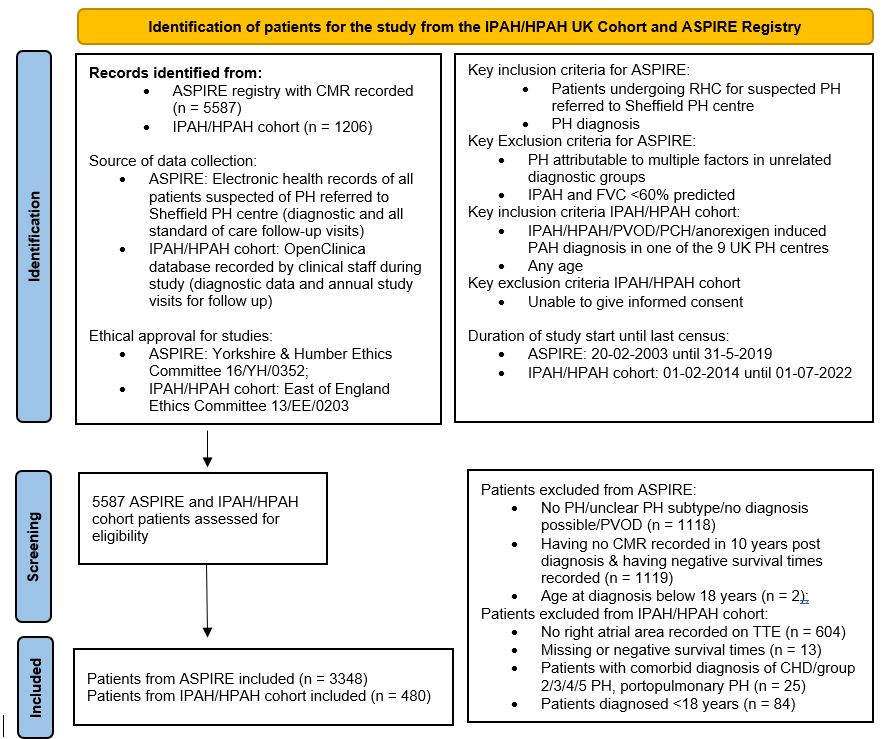

**Figure S1:** disposition of patients for the invasive and non-invasive mapping arm, National Cohort study and ASPIRE registry.

| Age/years | 53 ± 15 |
| --- | --- |
| Male sex | 19 (23.8) |
| Body mass index/kg.m^-2^ | 29.8 ± 5.6 |
| Genetic mutation  BMPR2  ALK1  TBX4 | 14 (17.5)  9 (11.3)  4 (5.0)  1 (1.3) |
| Time from diagnosis/years | 9.0 ± 5.0 |
| WHO Functional Class  I  II  III  IV | 5 (6.3)  30 (37.5)  37 (46.3)  8 (10.0) |
| Previous reported arrhythmia symptoms  Palpitations  Syncope/presyncope | 31 (38.8)  29 (36.3)  14 (17.5) |
| Prior ambulatory rhythm monitoring (source verified) | 12 (15.0) |
| Prior arrhythmia diagnosis  Atrial fibrillation (permanent)  Atrial tachycardia/flutter (paroxysmal)  Supraventricular tachycardia  Ventricular arrhythmia  Sinus node disease  2^nd^ or 3^rd^ degree AV block (intermittent) | 12 (15.0)  2 (2.5)  6 (7.5)  2 (2.5)  1 (1.3)  0  1 (1.3) |
| Baseline ECG  Atrial fibrillation/flutter  1^st^ degree AV block  Bundle branch block  1^st^ degree AV and bundle branch block  Frontal plane axis deviation | 4 (5.0)  4 (5.0)  9 (11.3)  3 (3.8)  30 (37.5) |
| Cardiac structure: MRI  RV end diastolic volume (indexed)/ml.m^-2^  RV end systolic volume (indexed)/ml.m^-2^  RV ejection fraction/%    LV end diastolic volume (indexed)/ml.m^-2^  LV end systolic volume (indexed)/ml.m^-2^  LV ejection fraction/%  RA area/cm^2^  RA volume (indexed)/ml.m^-2^  LA area/cm^2^ | N= 69  99.3 ± 36.7  59.1 ± 32.6  43.0 ± 13.8  71.7 ± 15.8  31.0 ± 9.1  63.3 ± 11.6  22.4 ± 6.0  39.9 ± 18.0  21.0 ± 5.3 |
| Pulmonary haemodynamics  Systolic PAP/mmHg  Diastolic PAP/mmHg  Mean PAP/mmHg  PCWP/mmHg  PVR (Thermodilution)/dynes  RA pressure/mmHg  Cardiac index  Vasoresponder | 82 ± 24  32 ± 11  50 ± 15  10 ± 3.5  899 ± 586  9 ± 5  2.2 ± 0.8  17 (21.3) |
| NT-proBNP/pgml^-1^ | 164 [76-786] |
| 6MW distance/metres | 370 ± 214 |
| Anti-arrhythmic therapy  Beta-blocker only  Amiodarone only  Non-DHP calcium channel blocker  Beta-blocker and amiodarone | 5 (6.3)  3 (3.8)  10 (12.5)  1 (1.3) |
| Vasodilator therapy  PDE5-inhibitor  Endothelin receptor antagonist  Selexipag  Intravenous prostanoid  Nebulised prostanoid  Riociguat  Calcium channel blocker  Single oral  Dual oral  Triple oral  Oral and parenteral  Intravenous only | 74 (92.5)  63 (78.8)  18 (22.5)  18 (22.5)  5 (6.3)  1 (1.3)  15 (18.8)  9 (11.3)  32 (40.0)  20 (25.0)  18 (22.5)  1 (1.3) |

**Table S1:** Baseline demographic and clinical characteristics of the study cohort at the time of loop recorder implant. Data presented as counts (%), mean ± standard deviation or median [interquartile range]. WHO= World Health Organisation; AV= atrioventricular; RV= right ventricular; RA= right atrial; LV= left ventricular; LA= left atrial; PAP= pulmonary artery pressure; PCWP pulmonary capillary wedge pressure=; PVR= pulmonary vascular resistance; 6MW= six-minute walk; DHP= dihydropyridine; PDE5= phosphodiesterase 5.

#### **Factors associated with arrhythmia incidence in non-invasive mapping cohort.**

On prospective follow-up cardiac arrhythmias were noted in 11 out of 30 (36.7%) of patients who underwent ECGi, some of whom suffered multiple different arrhythmias. These comprised atrial fibrillation (AF) (n=5), atrial tachycardia (n=2), sinus bradycardia (n=3), non-sustained ventricular tachycardia (nsVT) (n=1), and 2^nd^ or 3^rd^ degree atrioventricular block (n=2). 5 patients required specific treatment for their arrhythmia, while in 6 patients arrhythmia was silent and detected through ICM.

In arrhythmic patients ventricular activation-recovery interval was significantly shorter (294.6 ± 25.0 vs 267.3 ± 20.2ms, p=0.03). In keeping with our previous findings, MRI-derived RA area was larger in arrhythmic patients but this was not quite statistically significant (23.5 ± 5.0 vs 33.6 ± 21.9cm^2^, p=0.06). Atrial activation time was comparable between groups (74.6 ± 17.3 vs 95.1 ± 57.0ms, p=0.21), and there were no other significant differences in demographic, electrical or MRI-based parameters identified between patients with and without arrhythmia (table S6).

6 patients (4 arrhythmic, 2 non-arrhythmic) suffered clinical worsening events; 1 patient was hospitalised with right ventricular decompensation, 1 underwent lung transplantation, and 4 required uptitration of PH therapy due to disease progression. The risk of suffering a clinical worsening event if diagnosed with an arrhythmia was not statistically significant in this smaller cohort of patients (hazard ratio 3.5, 95% CI 0.8-15.9, p=0.11).

#### **Outcomes of patients requiring arrhythmia-specific intervention**

Of the 16 patients requiring changes to treatment as a consequence of significant arrhythmia: 2 were referred for ablation for SVT; 3 required uptitration or initiation of anti-arrhythmic drug therapy (including 2 requiring DC cardioversion) for SVT (n=2) or VT (n=1); 3 patients were newly diagnosed with atrial fibrillation, all asymptomatic, and required commencement of anticoagulation (total number of episodes = 6); 1 suffered an out of hospital ventricular fibrillation cardiac arrest resulting in hospitalisation and ultimately death; 2 had sustained ventricular tachycardia without haemodynamic compromise of whom 1 reported symptoms of palpitations; 2 required pacemaker insertion for high degree AV block, including 1 who was asymptomatic; 1 exhibited multiple sinus pauses of 2.5-6 seconds duration without symptoms; 2 had high degree atrioventricular block without symptoms including 1 with daytime episodes (figure S2).

|  | Comparator  N=71 | IPAH  N=80 | P val | Corr. P val |
| --- | --- | --- | --- | --- |
| Age/years | 52.7 ± 17.4 | 52.5 ± 14.9 | 0.94 | 0.98 |
| Male sex | 17 (23.9) | 19 (23.8) | 0.98 | 0.98 |
| Mean monitoring duration /days | 850 ± 253 | 848 ± 236 | 0.97 | 0.98 |
| Co-morbidity  Systemic hypertension  Diabetes mellitus  Ischaemic heart disease  Valvular heart disease  Hypercholesterolaemia | 11 (15.5)  3 (4.2)  2 (2.8)  2 (2.8)  8 (11.3) | 13 (16.3)  8 (10.0)  5 (6.3)  1 (1.3)  6 (7.5) | 0.90  0.17  0.32  0.49  0.43 | 0.98  0.62  0.85  0.87  0.87 |
| Arrhythmia  Tachycardia  AF  Atrial flutter/tachycardia/SVT  Ventricular arrhythmia  Bradycardia  Sinus node disease  2^nd^ or 3^rd^ degree AV block | 29 (40.8)  22 (31.0)  10 (14.1)  11 (15.5)  1 (1.4)  11 (15.5)  8 (11.3)  3 (4.2) | 32 (40.0)  23 (28.8)  4 (5.0)  16 (20.0)  5 (6.3)  11 (13.8)  5 (6.3)  7 (8.8) | 0.92  0.76  0.06  0.47  0.13  0.76  0.27  0.18 | 0.98  0.98  0.22  0.87  0.58  0.98  0.84  0.62 |

**Table S2:** Characteristics and arrhythmia frequency of study and comparator group. AF= atrial fibrillation; SVT= supraventricular tachycardia; AV= atrioventricular

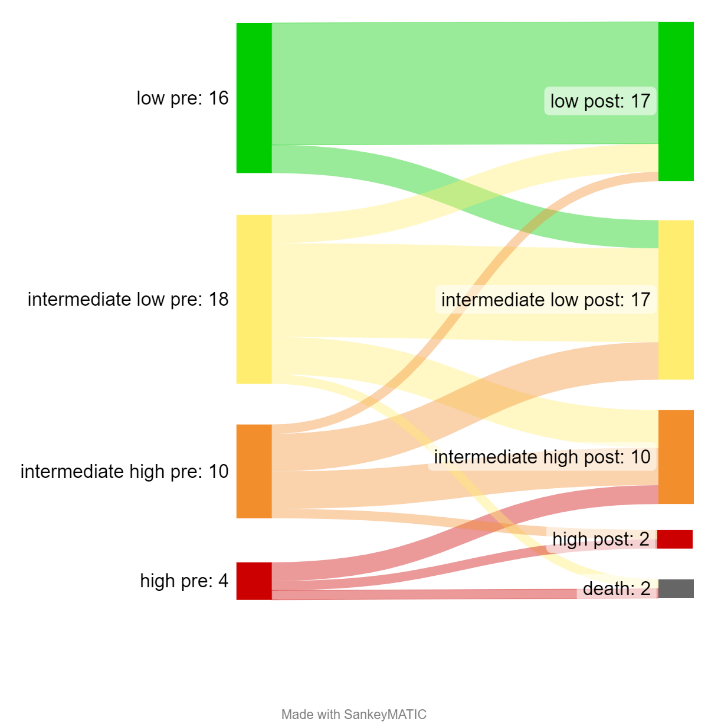

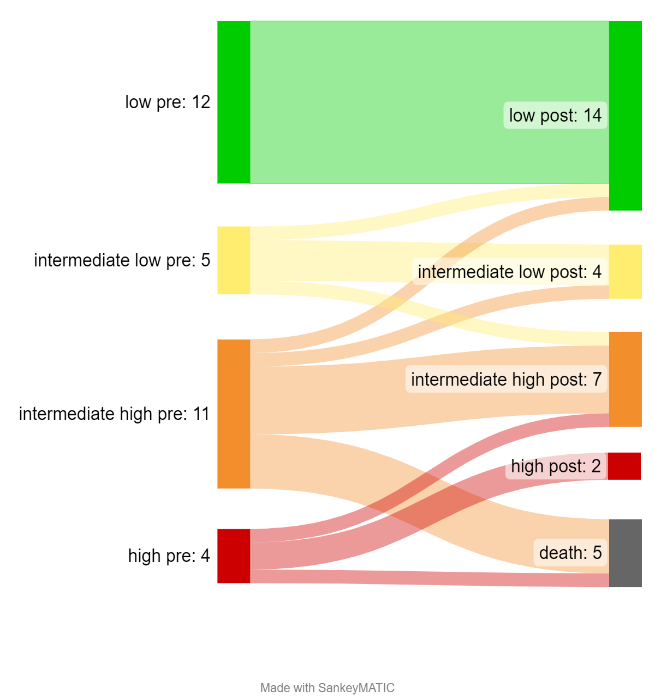

**B**
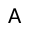
isk at baseline and at first follow-up and changes in risk**A**

**A**
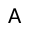
isk at baseline and at first follow-up and changes in risk**A**

**Figure S2:** Risk at time of ILR implant (‘pre’) and at end of study (‘post’), and changes in risk, as a function of presence or absence of arrhythmia. A= no arrhythmia, B= any arrhythmia

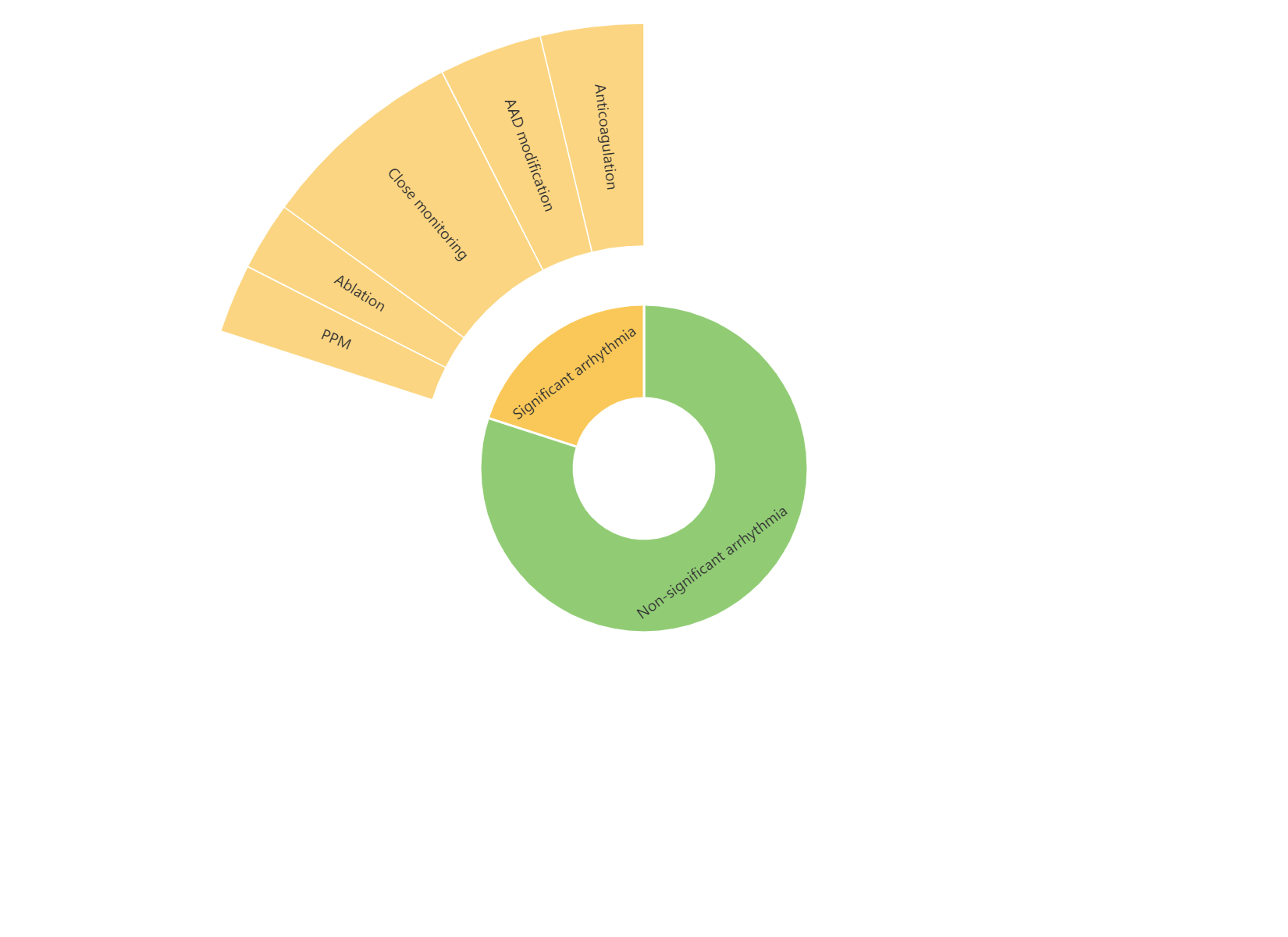

**Figure S3:** Proportions of patients experiencing clinically significant arrhythmia, and management of patients who did. AAD= anti-arrhythmic drug; PPM= permanent pacemaker.

|  | HR | 95% CI | P value |
| --- | --- | --- | --- |
| Arrhythmia symptoms | 1.31 | 0.44-2.33 | 0.25 |
| 1^st^ degree AV block or BBB | 2.27 | 0.42-3.37 | 0.19 |
| RA area | 1.03 | 1.01-1.08 | **0.005** |
| NTproBNP | 1.00 | 1.00-1.00 | 0.88 |

**Table S3:** Multivariable analysis of factors associated with significant arrhythmia occurrence. HR= hazard ratio; CI= confidence interval; AV= atrioventricular; BBB= bundle branch block; RA= right atrial)

|  | Invasive mapping  N=10 | Non-invasive mapping  N=30 |
| --- | --- | --- |
| Age/years | 63.0 ± 15.3 | 55.5 ± 13.2 |
| Male sex | 6 (60) | 11 (36.7) |
| BMI | 30.6 ± 7.8 | 28.0 ± 5.5 |
| WHO functional class  I  II  III  IV | 1 (10.0)  2 (20.0)  7 (70.0)  0 (0) | 7 (23.3)  13 (43.3)  10 (33.3)  0 (0) |
| Prior arrhythmia history | 10 (100) | 3 (10) |
| Pulmonary haemodynamics  mPAP/mmHg  PVR/dynes-sec.cm^5^  Cardiac Index/Lm^-2^  Mean RAP/mmHg  PAWP/mmHg | 49.5 ± 22.4  760 ± 602  2.3 ± 0.7  9.8 ± 5.4  12.2 ± 4.7 | 42.7 ± 10.5  672 ± 504  2.7 ± 0.8  7.2 ± 3.0  9.8 ± 2.7 |
| Pulmonary hypertension-specific therapy  No therapy  Monotherapy  PDE5i  ERA  Oral prostanoid  IV/SC/inh prostanoid  CCB  Dual therapy  PDE5i + ERA  PDE5i + oral prostanoid  PDE5i + IV/SC/inh prostanoid  PDE5i + CCB  ERA + oral prostanoid  Triple therapy  PDE5i + ERA + oral prostanoid  PDE5i + ERA + IV/SC/inh prostanoid | 2 (20.0)  1 (10.0)  0  0  0  0  4 (40.0)  0  0  0  0  3 (30)  0 | 0  1 (3.3)  1 (3.3)  0  0  2 (6.6)  9 (30.0)  1 (3.3)  0  1 (3.3)  2 (6.6)  11 (36.7)  2 (6.6) |
| 6-minute walk distance/m | 367.0 ± 162.9  *(n=9)* | 425.1 ± 174.7 |
| NT pro-BNP/pgml^-1^ | 961 [485-1493] | 204 [81-375] |

**Table S4:** Demographic, anatomical, haemodynamic and functional characteristics of the invasive and non-invasive mapping cohorts. BMI= body mass index; WHO= World Health Organisation; mPAP= mean pulmonary arterial pressure; PVR= pulmonary vascular resistance; RAP= right atrial pressure; PAWP= pulmonary arterial wedge pressure; PDE5i= phosphodiesterase 5 inhibitor; ERA= endothelin receptor antagonist; IV= intravenous; SC= subcutaneous; inh= inhaled; CCB= calcium channel blocker.

| **Pt no.** | **Age** | **Sex** | **PAH group** | **Clinical tachycardia(s)** | **Complications** | **Follow-up** | **RA vol./ml** | **AVNERP/ms** | **PH-related CWE** |
| --- | --- | --- | --- | --- | --- | --- | --- | --- | --- |
| 1 | 40-45 | M | 4- CTEPH | Typical flutter | nil | No recurrence at 12 months off anti-arrhythmics | 255 | 320 | Nil |
| 2 | 76-80 | F | 4- CTEPH | Typical flutter | nil | No flutter recurrence. AF 3 weeks post-procedure, requiring reintroduction of amiodarone. No further atrial arrhythmia by 12 months. | 261 | 330 | Death 24 months post-ablation |
| 3 | 76-80 | M | 4- CTEPH | Atypical flutter- lateral RA free wall | nil | AF noted at first follow-up. Death 9 months post-procedure. | 287 |  | Death 21 months post-ablation |
| 4 | 36-40 | M | 1.2- HPAH | Typical flutter | nil | No recurrence at 12 months off anti-arrhythmics | 368 | 550 | Nil |
| 5 | 40-45 | M | 1.4- a/w CHD (closed VSD) | Typical flutter | nil | No flutter recurrence. AF 2 weeks post-procedure requiring cardioversion and reinitiation of amiodarone. No further atrial arrhythmia by 12 months | 398 | 530 | Clinical deterioration requiring therapy uptitration 21 months post-ablation |
| 6 | 60-65 | F | IPAH | Typical flutter | nil | No recurrence at 12 months off anti-arrhythmics | 179 | 350 | Nil |
| 7 | 70-75 | F | 1.4- a/w CHD (ASD) | Typical flutter; AF | nil | No recurrence at 12 months off anti-arrhythmics | 167 |  | Nil |
| 8 | 70-75 | M | 4- CTEPH | Typical flutter; focal AT | Junctional brady (slow sinus node recovery): TPW placed, monitored. | No recurrence at 6 months off anti-arrhythmics | 300 | 230 | Nil |
| 9 | 66-70 | M | 4- CTEPH | Typical flutter | nil | No recurrence at 6 months off anti-arrhythmics | 207 | 400 | Nil |
| 10 | 70-75 | F | 4- CTEPH | Atypical clockwise CTI-dependent flutter | nil | No recurrence at 6 months off anti-arrhythmics | 256 | 440 | Nil |

**Table S5:** demographic and procedural characteristics of patients undergoing invasive mapping and ablation for atrial arrhythmia.

|  | Lean  N=16 | Obese  N=16 | PAH  N=16 | P val |
| --- | --- | --- | --- | --- |
| Age/years | 42 ± 11 | 43 ± 12 | 47 ± 9 | 0.39 |
| Male sex | 3 (18.8) | 3 (18.8) | 3 (18.8) | 1.00 |
| BMI | 22.8 ± 2.6 | 46.7 ± 5.5 | 27.9 ± 6.1 | <0.0001 |
| Conduction properties  Atrial activation time/ms  Vent. activation time/ms  Vent. repolarisation time/ms  Vent. ARIc/ms | 46 ± 12  31 ± 6  87 ± 25  246 ± 25 | 62 ± 15  34 ± 5  105 ± 16  247 ± 19 | 72 ± 17  41 ± 13  127 ± 51  292 ± 30 | <0.0001  0.013  0.010  <0.0001 |

**Table S6:** Clinical and cardiac electrophysiological, anatomical and functional features associated with arrhythmia. BMI= body mass index

R=0.29 p=0.20

RV EDVi/ml.m-2

| RV T1 | MRI_RVEDVi |
| --- | --- |
| 1005.5 | 120 |
| 1104.8 | 98 |
|  | 92 |
| 1204.5 | 108 |
| 1154.3 | 98 |
| 1045.5 | 58 |
| 1294.8 | 111 |
|  | 98 |
| 1194.8 | 84 |
|  | 102 |
| 1308.3 | 102 |
| 1268.8 | 96 |
| 1395 | 117 |
| 1310 | 91 |
|  | 179 |
| 1331.5 | 149 |
| 1353.5 | 104 |
| 1243.5 | 148 |
| 1321.5 | 55 |
|  | 151 |
| 1301.3 | 116 |
| 1019.3 | 68 |
|  | 56 |
| 1212.8 | 71 |
|  | 91 |
|  | 137 |
| 1249.5 | 138 |
| 1277.5 | 66 |
|  | 161 |
| 1168.8 | 80 |

R=-0.005 p=0.99

VAT/ms

| RV T1 | Total activation time (95% of all data) |
| --- | --- |
| 1005.5 | 45.4102 |
|  | 45.4102 |
| 1204.5 | 35.6445 |
| 1154.3 | 44.4336 |
| 1294.8 | 33.6914 |
| 1194.8 | 84.4727 |
|  | 25.3906 |
| 1308.3 | 70.3125 |
| 1395 | 39.0625 |
|  | 30.7617 |
| 1331.5 | 54.6875 |
| 1353.5 | 19.5312 |
| 1243.5 | 36.6211 |
| 1321.5 | 36.1328 |
| 1019.3 | 30.2734 |
| 1212.8 | 28.3203 |
|  | 40.0391 |
| 1249.5 | 97.6562 |
| 1277.5 | 36.1328 |
|  | 63.9648 |

**Figure S4:** Relationship between right ventricular scar burden, assessed by T1 mapping, and (left) right ventricular end diastolic volume (indexed to body surface area) and (right) ventricular activation time. RV= right ventricle; EDVi= indexed end diastolic volume; VAT= ventricular activation time.

R=-0.58 p=0.006

RV EF/%

| RV T1 | MRI_RVEF |
| --- | --- |
| 1005.5 | 62 |
| 1104.8 | 56 |
|  | 58 |
| 1204.5 | 52 |
| 1154.3 | 45 |
| 1045.5 | 53 |
| 1294.8 | 34 |
|  | 53 |
| 1194.8 | 58 |
|  | 37 |
| 1308.3 | 33 |
| 1268.8 | 38 |
| 1395 | 45 |
| 1310 | 43 |
|  | 32 |
| 1331.5 | 47 |
| 1353.5 | 55 |
| 1243.5 | 35 |
| 1321.5 | 52 |
|  | 56 |
| 1301.3 | 40 |
| 1019.3 | 61 |
|  | 40 |
| 1212.8 | 51 |
|  | 59 |
|  | 33 |
| 1249.5 | 36 |
| 1277.5 | 50 |
|  | 24 |
| 1168.8 | 53 |

Rad.: R=0.14, p=0.56 Circ.: R=0.12, p=0.61 Long.: R=0.51, p=0.04

RV strain

| RV T1 | RV strain radial | RV strain circumferential | RV strain longitudinal |
| --- | --- | --- | --- |
|  | -5 | -3.56 | -18.44 |
| 1104.8 | -17.8 | -12.15 | -24.09 |
|  | -15.2 | -10.76 | -21.29 |
| 1204.5 | -13.4 | -9.93 |  |
| 1154.3 | -20 | -13.84 |  |
| 1045.5 | -18.1 | -11.82 | -24.89 |
| 1294.8 | -8.2 | -6.51 | -18.88 |
| 1194.8 | -13.2 | -8.93 | -28.01 |
|  | -16.3 | -11.9 | -20.93 |
| 1308.3 | -16.8 | -11.87 | -13.58 |
| 1268.8 | -11.1 | -8.14 | -14.65 |
| 1395 | -18.7 | -12.3 | -20.26 |
| 1310 | -16.6 | -11.65 | -18.32 |
|  | -11.2 | -8.19 | -11.13 |
| 1331.5 | -16.7 | -11.59 | -17.27 |
| 1353.5 | -18.1 | -12.43 | -21.2 |
| 1243.5 | -9.6 | -7.44 | -10.76 |
|  | -30.1 | -17.83 | -25.25 |
| 1301.3 | -15.2 | -10.99 | -19.02 |
| 1019.3 | -16.9 | -11.52 | -23.32 |
|  | -12.3 | -8.56 |  |
| 1212.8 | -23.7 | -14.94 | -26.18 |
|  | -20 | -12.85 | -21.65 |
|  | -9.4 | -7.67 | -14.16 |
| 1249.5 | -14.4 | -10.38 | -17.46 |
| 1277.5 | -21.2 | -13.38 |  |
| 1168.8 | -19.3 | -12.85 | -23 |

**Figure S5:** Relationship between right ventricular scar burden, assessed by T1 mapping, and (left) right ventricular ejection fraction and (right) strain. RV= right ventricle; EF= ejection fraction.

|  | No arrhythmia  N=19 | Arrhythmia  N=11 | P value |
| --- | --- | --- | --- |
| Age | 54.1 ± 13.0 | 57.9 ± 13.7 | 0.45 |
| Male sex | 5 (26.3) | 6 (54.5) | 0.24 |
| BMI | 28.1 ± 6.2 | 27.8 ± 4.2 | 0.91 |
| Electrophysiologial parameters  Vent. activation time/ms  Vent. repolarisation time/ms  Vent. activation recovery interval/ms    Atrial activation time/ms | 43.0 ± 17.3  114.2 ± 34.3  294.6 ± 25.0  *N=14*  74.6 ± 17.3  *N=15* | 49.4 ± 26.9  150.9 ± 45.4  267.3 ± 20.2  *N=6*  95.1 ± 57.0  *N=9* | 0.53  0.06  0.03  0.21 |
| MRI functional parameters  LV strain  radial  circumferential  longitudinal  LV ejection fraction  RV strain  radial  circumferential  longitudinal  RV ejection fraction  RV T1 mapping  Posterior  Free wall  Anterior  Septal  Average | -30.0 ± 6.7  -18.0 ± 2.7  -16.4 ± 3.0  60.4 ± 8.5  -15.3 ± 5.5  -10.6 ± 3.2  -19.3 ± 5.0  46.7 ± 9.6  1232 ± 115  1220 ± 147  1297 ± 161  1193 ± 126  1236 ± 116 | -27.0 ± 7.0  -16.7 ± 3.1  -15.8 ± 3.0  54.6 ± 8.9  -16.9 ± 4.6  -11.5 ± 2.3  -20.7 ± 4.1  45.7 ± 11.9  1152 ± 149  1175 ± 91  1278 ± 174  1214 ± 170  1205 ± 102 | 0.29  0.31  0.64  0.09  0.46  0.48  0.53  0.80  0.20  0.50  0.81  0.76  0.58 |
| MRI anatomical parameters  LV EDV(i)  LV ESV(i)  LV SV  RV EDV(i)  RV ESV(i)  RV SV    RA area  LA area | 70.7 ± 11.8  28.6 ± 8.9  76.9 ± 14.8  105.8 ± 29.3  58.5 ± 24.0  83.8 ± 22.6  23.5 ± 5.0  21.0 ± 3.8 | 67.6 ± 19.2  29.5 ± 9.1  70.6 ± 22.0  103.2 ± 38.4  58.0 ± 31.7  85.3 ± 25.1  33.6 ± 21.9  22.0 ± 6.0 | 0.59  0.80  0.36  0.84  0.96  0.87  0.06  0.56 |
| Clinical worsening event | 2 (10.5) | 4 (36.3) | 0.09 |

**Table S7:** Clinical and cardiac electrophysiological, anatomical and functional features associated with arrhythmia in the non-invasive mapping group. BMI= body mass index; LV= left ventricle; RV= right ventricle; EDV= end diastolic volume; ESV= end systolic volume; SV= stroke volume; RA= right atrium; LA= left atrium.

|  | HR | 95% CI | P value |
| --- | --- | --- | --- |
| RA area <22.8cm^2^ | 0.59 | 0.38-0.91 | 0.017 |
| Male sex | 1.89 | 1.22-2.93 | 0.005 |
| Age | 1.06 | 1.04-1.07 | <0.001 |
| RA pressure | 1.00 | 0.95-1.04 | 0.90 |
| mPAP | 1.02 | 0.99-1.05 | 0.29 |
| PVR | 1.00 | 1.00-1.00 | 0.20 |
| Cardiac output | 0.66 | 0.48-0.90 | 0.009 |

**Table S8:** multivariable analysis of clinical factors associated with mortality in patients with group 1 pulmonary arterial hypertension. RA= right atrium; mPAP= mean pulmonary artery pressure; PVR= pulmonary vascular resistance; HR= hazard ration; CI= confidence interval

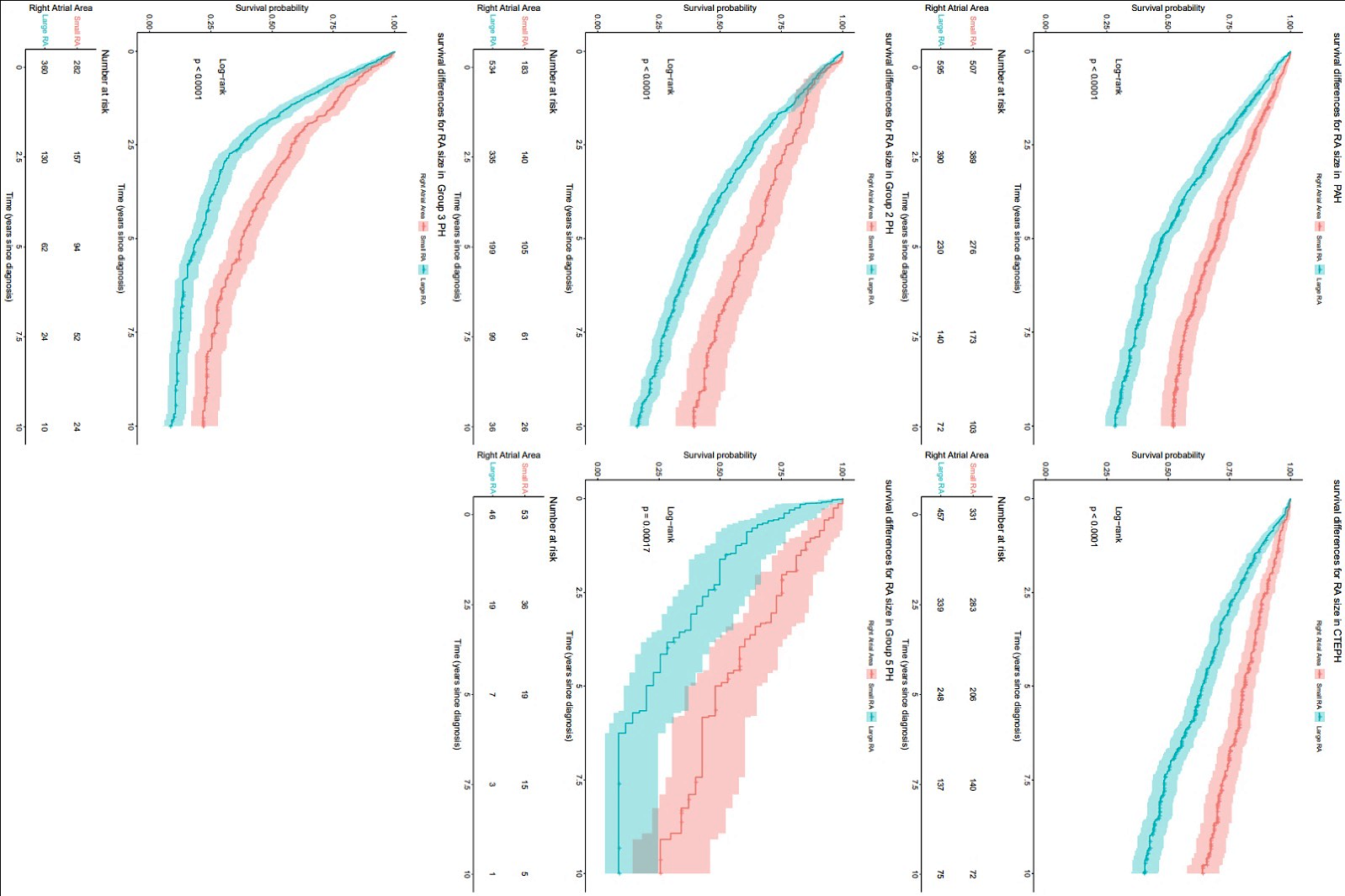

**Figure S6:** Kaplan-Meier survival curves for all pulmonary hypertension groups as a function of right atrial size.
